# The Effects of Allowing Professional Incorporation on Physician Labour Supply

**DOI:** 10.1101/2025.03.14.25323936

**Authors:** David Kanter-Eivin, Helena Son, Adam Steiner, Stephenson Strobel

## Abstract

We study how tax deferral policies shape labour supply decisions over the lifecycle. Between 1975 and 2005, Canadian provinces allowed physicians to incorporate, providing high-income professionals with substantial tax deferral opportunities. Using a difference-in-differences identification strategy, we find that incorporation reduced work effort by facilitating earlier retirement. Physician supply fell by 6.8% over the long run, with especially large declines among older physicians and high-earning surgical specialists. These results provide long-run evidence of how these tax deferral policies, designed to affect high-skill labour, can alter timing of retirement and reduce labour supply.

## 1 Introduction

Tax incentives are among the most widely used policy tools for addressing professional labour shortages, yet their effectiveness remains theoretically ambiguous and empirically understudied. While substitution effects suggest that improving the returns to work should increase labour supply, income effects may operate in the opposite direction when policies effectively increase lifetime wealth. This tension is particularly relevant for high-skilled professionals, who face both steep tax schedules and complex intertemporal career decisions. Untangling the relative importance of these mechanisms is central to understanding life-cycle labour supply and retirement behaviour, particularly among high-income professionals with substantial autonomy over their work and savings decisions.

We study this question using a natural experiment in professional tax policy. Between 1975 and 2005, Canadian provinces sequentially allowed physicians to professionally incorporate, creating substantial tax deferral benefits worth thousands of dollars annually. Professional incorporation permits physicians to retain earnings within their corporation and defer personal income taxes until withdrawal, typically during retirement when they face lower marginal tax rates. This policy was explicitly designed to address physician shortages by improving the financial incentives for medical practice. Physician advocacy groups lobby heavily for professional incorporation, suggesting a degree of revealed preference (Ontario Medical Association, 2024; Canadian Medical Association, 2024; Ross, 2024; Herrling, 2017). Lower-bound estimates are that physicians in Canada can reduce their annual tax burden by 4% and retain savings within a corporation of 10,000 per year (Nielsen and Sweetman, 2018).

The staggered provincial adoption of incorporation creates an ideal setting to identify causal effects of tax deferral policies on professional labour supply. Economic theory provides competing predictions: incorporation may increase current work effort as physicians substitute toward periods with higher effective returns (substitution effect), or it may reduce work effort by increasing lifetime wealth and facilitating earlier retirement (income effect). The net effect depends on the relative magnitude of these opposing forces and may vary systematically with income levels, career stage, and other professional characteristics.

We first establish evidence on how in-demand professional incorporation is by describing the adoption of professional incorporation in the provinces of Ontario and Manitoba. We use scraped College of Physicians and Surgeons of Ontario (CPSO) and College of Physicians and Surgeons of Manitoba (CPSM) licensing data. These data include all physicians who have ever been registered in the province of Ontario from 1940 until 2024 and all current physicians in the province of Manitoba as of 2025. Since it is mandatory for incorporated physicians to register their professional corporation with the physician regulatory bodies, we are able to observe the prevalence of physicians who incorporate.

We find that after 2003, when CPSO regulations allowed incorporation, there was an explosion of professional incorporation such that 40% of all physicians ever registered in Ontario incorporated. Of active practicing physicians in 2024, 68% have incorporated. 61% of all actively practicing physicians also incorporate in Manitoba. At least in these two provinces, most physicians find it in their best interest to incorporate.

We then answer whether allowing incorporation meaningfully affects intensive and extensive margin supply of physician labour. Using a difference-in-differences analysis that compares provinces before and after allowing incorporation, we find that income effects dominate substitution effects. Rather than increasing physician labour supply, allowing physician incorporation reduces both intensive and extensive margin supply in Canada. We find reductions in intensive margin labour supply among surgical specialists by 13% over our period of observation. This especially reduces the provision of procedures among this group of physicians.

Five years after implementation, extensive physician supply also falls by 1.9%, with larger effects among surgical specialists and older physicians. These extensive margin effects persist and strengthen over time, with significant reductions still evident twenty years post-adoption. Over the long run of 23 years in our data, total physician supply falls by 6.8% with impacts on both medical and surgical specialists.

The heterogeneity in our results supports the theoretical prediction that income effects should be strongest among high earners who benefit most from tax deferral. Surgical specialists, who earn significantly more than family physicians, show the largest negative responses in both service provision and labour force participation. Effects are also concentrated among physicians over age 60 and those more than 30 years from graduation. These are the groups most likely to be considering retirement and thus most responsive to policies that improve retirement financing. Consistent with the policy affecting retirement, the age structure of the physician labour force in Canada changes in response such that average age of physicians decreases by 0.7 years and the median age decreases by 1.1 years.

Our contribution is threefold. First, we provide novel evidence on intertemporal labour supply elasticities among high-income professionals, complementing existing work on retirement incentives and social insurance. Second, we show that tax policies designed to increase professional retention may unintentionally reduce labour supply by facilitating earlier workforce exit. Third, we demonstrate heterogeneity consistent with life-cycle models: effects are concentrated among physicians nearing retirement and among specialties that benefit most from tax deferral. Together, these results highlight how policies that shift lifetime wealth without raising current marginal returns to work can meaningfully affect labour supply through altered retirement timing.

Our findings underscore a broader policy insight: when tax policies primarily increase lifetime wealth rather than current marginal returns to work, they may unintentionally reduce labour supply. This has implications beyond healthcare, as similar incorporation and deferral schemes exist across other professional sectors (Kopczuk and Zwick, 2020). By focusing on a high-income, high-autonomy workforce with staggered access to tax deferral, we provide rare empirical evidence that tax design can shape when, not just whether, professionals choose to work.

As populations age and demand for professional services grows, understanding these effects becomes crucial for effective policy design. More broadly, our findings speak to fundamental questions in public finance about the effectiveness of tax incentives in changing behaviour versus simply subsidizing existing plans. The large, persistent negative effects we document suggest that policymakers should carefully consider the intertemporal incentives created by tax policies, particularly when targeting high-income professionals who have both the resources and flexibility to respond to such incentives through altered retirement timing.

### 1.1 Institutional details of physician incorporation in Canada

Professional incorporation for physicians was first permitted in Alberta in December 1975. The policy aimed to allay physician concerns about the potential impact of recently implemented universal health care on their incomes. Contemporary analyses estimated that incorporation allowed physicians to defer a tax burden equivalent to 12% of annual income. Within two years, more than 20% of physicians in the province had incorporated (Gray, 1978; Imlach, 1976).

Other provinces began allowing professional incorporation several years after Alberta (Government of New Brunswick, 1981; Government of Quebec, 2001; Government of Manitoba, 1999; Government of British Columbia, 1990) with similar policy rationales. Allowing incorporation was intended to retain physicians by boosting their effective incomes (Doctor’s Nova Scotia, 2017). Saskatchewan (College of Physicians and Surgeons of Saskatchewan, 2017) and New Brunswick (Government of Nova Scotia, 1996) subsequently allowed incorporation in 1980 and the final province, Newfoundland and Labrador, allowed professional incorporation in 2005 (Government of Newfoundland and Labrador, 2005). Ontario, which legislated incorporation in 2001 (Ontario Medical Association Legal Services, 2008), allowed physicians to incorporate through regulation in 2003. Similar to Alberta, the prevalence of incorporation of physicians increased was 54% five years after it was allowed in Ontario (Nielsen and Sweetman, 2018). Table 1 shows the year of legislation within each province.

**Table 1:** Years that provinces legislated professional incorporation. Note that this legislation is earlier than when a physician can effectively incorporate as licensing colleges must pass regulations for this purpose.

| Province | Year of Adoption |
| --- | --- |
| Alberta | 1975 |
| Saskatchewan | 1981 |
| New Brunswick | 1981 |
| Prince Edward Island | 1988 |
| British Columbia | 1990 |
| Nova Scotia | 1996 |
| Manitoba | 1999 |
| Ontario | 2000 |
| Quebec | 2001 |
| Newfoundland and Labrador | 2005 |

Professional incorporation offers several advantages to Canadian physicians. Physicians may draw salaries as employees of their corporations, with such payments taxed at the applicable personal income tax (PIT) rate. By contrast, income retained within a professional corporation is subject to the lower corporate income tax (CIT) rate and can be used to purchase investment assets: a conservative estimate places retained income at $10,000–$15,000 per year (Nielsen and Sweetman, 2018). Corporate earnings may also be used for immediate expenses, such as paying contractors or employees other than the physician. These are often family members providing services like bookkeeping. Until federal tax reforms in 2018, family members could also hold non-controlling shares in a professional corporation and receive dividends, enabling income splitting when those family members faced lower marginal tax rates (Wolfson et al., 2016).

However, professional incorporation does not confer all the advantages associated with more typical forms of incorporation. In particular, it does not shield physicians from liability related to medical practice. Moreover, the sale of medical practices is uncommon in Canada, so the value of a professional corporation is often limited to its investment assets.

The tax advantages of incorporation can be understood by comparing long-run differences between personal and corporate tax rates. Figure 1 illustrates how small business corporate income tax (CIT) rates—combining federal and provincial components—have evolved relative to top marginal personal income tax (PIT) rates across provinces over the past 50 years (Finances of the Nation, 2025). In the 1970s, PIT rates were extremely high, often ranging from 70 to 90%. Although lower today, Canadian PIT rates remain elevated compared to many peer countries. For example, Ontario’s combined top marginal PIT rate in 2022 was 53%. By contrast, CIT rates have consistently been far lower. The highest observed CIT rate during the sample period was 39% in Newfoundland and Labrador in 1977, while Ontario’s 2022 CIT rate was only 12.2%. In every province and year, the top marginal PIT has exceeded the corresponding CIT by a wide margin.

**Figure 1:**
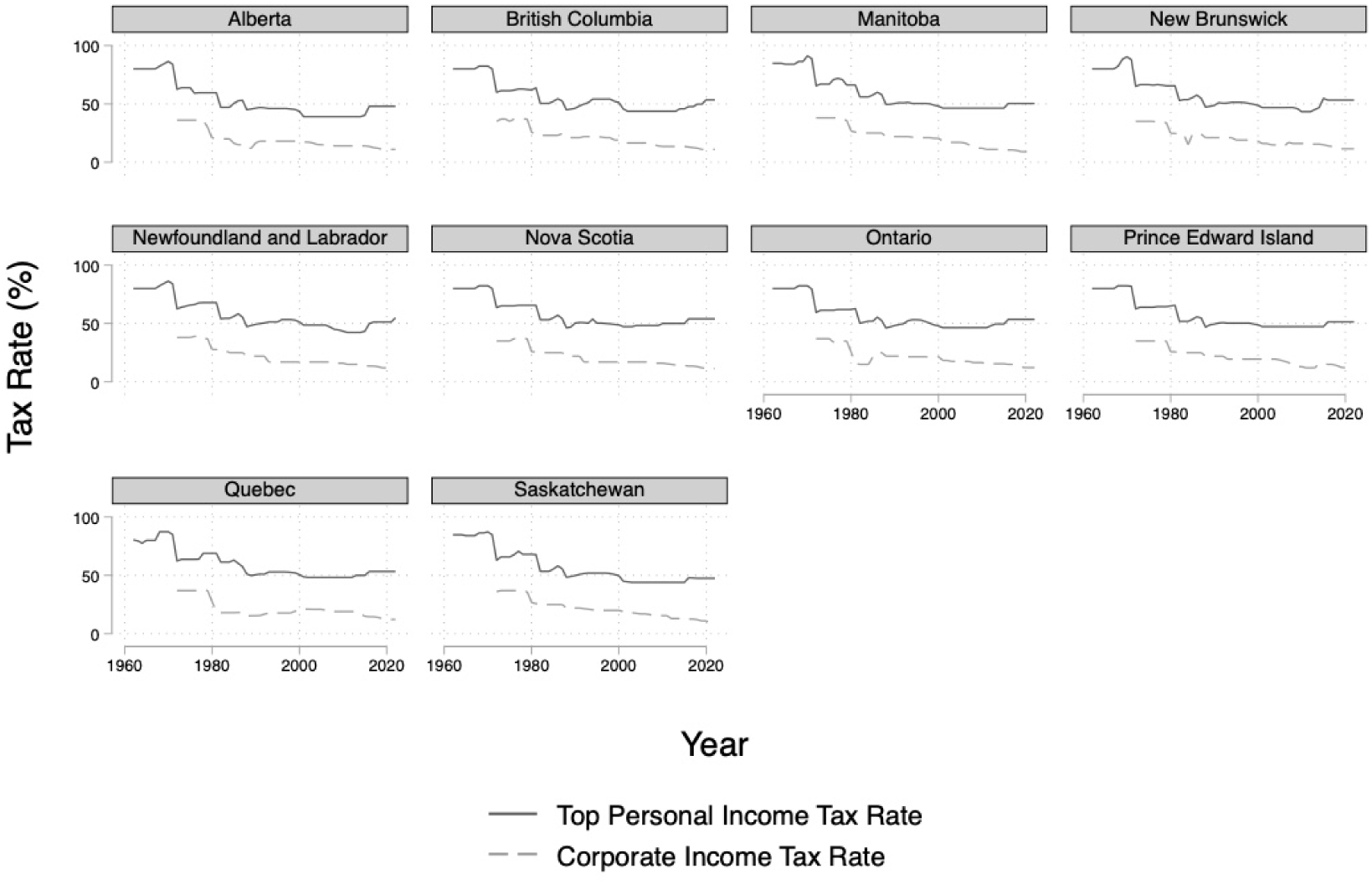
Total small business (comprising provincial and federal) tax rates and top personal income tax rates over the period of 1970 to 2020.

### 1.2 Previous Literature

Our research contributes to four main strands of literature: (1) financial incentives and physician labour supply; (2) intertemporal labor-leisure substitution; (3) retirement behavior and social insurance; and, (4) income effects among high-earning individuals.

A large literature studies how income and wage changes affect physician labor supply, generally finding modest responses. Estimated elasticities vary substantially: Sloan (1975), Hickson et al. (1987), and Devlin and Sarma (2008) document small or moderate effects, and policy simulations by Kalb et al. (2018) suggest that a 1% wage increase leads to only a 0.1–0.5% increase in hours worked. Similar inelastic responses have been found in Australia (McRae and Butler, 2014), while American settings reveal greater elasticity (Yang, 1987; Clemens and Gottlieb, 2014). Some studies decompose income and substitution effects: Rizzo and Blumenthal (1994) estimate that while substitution effects are positive, pure income effects may actually reduce labor supply. Notably, evidence also suggests that physicians adjust their retirement decisions in response to income shocks, even after peak earnings years (Gottlieb et al., 2025). Our setting differs in that incorporation does not immediately increase income, but instead creates the option to defer income into retirement, producing a delayed wealth effect that can influence retirement timing.

Our research also contributes to a broader literature on labor-leisure tradeoffs and intertemporal responses to financial incentives. Classic life-cycle models posit that workers smooth consumption over time in response to permanent income changes (Friedman, 1957; Lucas and Rapping, 1969). Early empirical estimates, however, suggest limited intertemporal elasticity of labor supply (Altonji, 1982). More recent work using windfall income finds persistent reductions in earnings from lottery winnings (Cesarini et al., 2017), consistent with long-run consumption smoothing. In the context of professional incorporation, we interpret the tax-deferral mechanism as a delayed wealth shock that may trigger earlier retirement and reduced late-career labour supply.

Evidence from other social policy interventions suggests that labour responses to income transfers can vary depending on the design and targeting of programs. In Canada, child tax benefits did not meaningfully affect labour supply (Baker et al., 2023), nor did the removal of work requirements from the U.S. Child Tax Credit (Enriquez et al., 2023). However, programs explicitly tied to work, such as subsidized childcare in Quebec (Baker et al., 2008) and the U.S. Earned Income Tax Credit (Bastian and Lochner, 2022), have been shown to increase labour force participation. Other studies also document reductions in labour supply in response to more generous unemployment insurance, as in Canada during the 1980s (Card and Freeman, 1993). These examples emphasize that behavioural responses are strongest when incentives are tightly linked to labour market participation. This is true for incorporation, which conditions the full tax benefit on reduced income from work.

Our paper also contributes to literature on retirement decisions and social security policy. Much of this work explores how changes in the level or rules of public pension systems influence labour force participation. Krueger and Pischke (1992), Burtless (1986), and Blau and Goodstein (2010) find only modest effects of social security generosity on retirement. In contrast, simulation studies suggest that benefit cuts or changes to eligibility age can meaningfully delay retirement (French, 2005; Coile and Gruber, 2007; Mastrobuoni, 2009). Institutional rule changes, such as raising the early retirement age, have been shown to increase participation among older workers (Staubli and Zweimüller, 2013), and some scholars argue that these rules — rather than benefit levels — explain much of the labour supply response (Rust and Phelan, 1997). In this spirit, we interpret incorporation as a rule-based policy change that lowers the cost of retirement by facilitating pre-tax savings, thereby increasing the attractiveness of early labour force exit.

Finally, we situate this paper within an empirical literature on the labour supply and retirement behaviour of high-income earners. Pass through businesses like the incorporation we study have become more prevalent in the US and international settings. This has implications for inequality (Kopczuk and Zwick, 2020). In Canada, physicians make up a substantial share of top earners despite recent declines in relative income rank (Riddell and Lemieux, 2015). Higher-income individuals tend to save more, although not always for retirement purposes (Dynan et al., 2004), and may respond differently to tax incentives. For example, tax holidays in Switzerland disproportionately reduced labour supply among wealthier individuals (Martínez et al., 2021). In Canada, higher-income workers may be less likely to retire at conventional ages like 60 or 65, and appear to place more weight on non-pecuniary factors in deciding when to exit the workforce (Milligan and Schirle, 2024). Some evidence also suggests that accrual of “income-security wealth”, such as pensions, has offsetting effects on retirement: it can both delay exit by raising opportunity cost and accelerate exit by increasing financial readiness (Baker et al., 2001; Schirle, 2010). Our results suggest that incorporation facilitates this latter mechanism: it allows physicians to accumulate wealth in a tax-advantaged way, thereby enabling earlier retirement.

## 2 Theoretical Framework

To understand the mechanisms through which professional incorporation affects physician labour supply, we develop a two-period model where physicians make intertemporal consumption and labour supply decisions. The details of this model are in Appendix B. The model incorporates key features of the Canadian tax system and physician incorporation rules to generate testable predictions about both intensive and extensive margin responses.

### 2.1 Model Setup

Our theoretical framework models physicians as utility-maximizing agents who live for two periods and choose labour supply and consumption in each period. Period 1 represents prime working years, while period 2 represents the pre-retirement period where physicians decide whether to continue working or retire early. Physicians have Stone– Geary preferences with subsistence levels for both consumption and leisure, creating potential threshold effects in labour supply decisions.

The key innovation of professional incorporation is that it fundamentally alters physicians’ budget constraints in two ways. First, incorporation allows physicians to save from pre-tax rather than after-tax income, effectively increasing the return to deferred consumption in after-tax terms. Second, incorporation provides enhanced flexibility in tax timing strategies, as physicians can control their taxable income in period 2 through both labour supply choices and accumulated savings withdrawal decisions. Canada has a progressive tax structure, which creates incentives for incorporated physicians to strategically manage later incomes to potentially qualify for the lower tax rate. If a physician can limit their later life income, they can then pay themselves out of the savings they hold within their corporation at a lower income tax rate.

### 2.2 Key Economic Mechanisms

The model identifies competing forces that determine the net effect of incorporation on labour supply. Substitution effects operate through the increased effective return to saving, which may encourage greater work effort in earlier periods to fund higher consumption or facilitate earlier retirement. Income effects operate through the overall improvement in the physician’s budget constraint, making them effectively wealthier and potentially incentivizing more leisure.

The presence of subsistence consumption and leisure thresholds in the Stone–Geary utility function creates the possibility of threshold effects and corner solutions. If a physician’s accumulated savings under incorporation are sufficient to meet subsistence consumption needs in later periods, the optimal response may be complete labour force exit. This generates the potential for discontinuous, extensive-margin responses such as early retirement.

### 2.3 Theoretical Predictions

Our model generates several empirically testable predictions:

**Dominance of Incorporation**: Under the assumption that incorporation is costless, incorporation weakly dominates non-incorporation for all physician types due to the expanded choice set it provides.

**Ambiguous Intensive Margin Effects**: The effect of allowing incorporation on current work effort (period 1 labour supply) is theoretically ambiguous. While the higher effective return to savings creates substitution effects that may increase work effort, the resulting increase in lifetime wealth produces income effects that may reduce effort. Both positive and negative intensive margin responses are feasible.

**Negative Extensive Margin Effects**: Incorporation creates strong incentives for early retirement by enabling physicians to defer taxes on savings and smooth consumption in retirement. We expect to observe declines in extensive margin labour supply, particularly among older physicians and those approaching traditional retirement ages.

**Heterogeneous Responses**: Since physicians with sufficient earnings can meaningfully benefit from tax deferral, we expect stronger responses among high-earning specialties and more experienced physicians who have accumulated greater savings over their careers.

The model’s emphasis on threshold effects and the discrete nature of retirement decisions suggests that incorporation may trigger sharp behavioural responses, including abrupt transitions from full-time practice to complete retirement, rather than gradual reductions in work effort. These theoretical predictions guide our empirical analysis, which examines both short-run and long-run changes in physician labour supply at the intensive and extensive margin. We pay particular focus on how effects vary by physician specialty, age, and career stage. These are the dimensions along which our model predicts the strongest heterogeneous responses.

## 3 Methods

### 3.1 Data

We use three sources of data to evaluate effects of incorporation on intensive and extensive margin physician supply. First, to establish a first stage effect on uptake we examine physician incorporation in Ontario and Manitoba. We use a unique set of data from the CPSO’s “Doctor Search” and CPSM’s “Registrants’ Directory” websites. These data were scraped in 2024 and 2025, respectively. The CPSO data contain all current and former physicians licensed in the province of Ontario from the 1940s to 2024. The CPSM data contains all actively practicing physicians as of 2025. Both registries contain information on physician characteristics like specialty, year of graduation, medical school of graduation, year of registration with the CPSO and CPSM, and whether the physician is incorporated. As all physicians in Canada must register with their provincial licensing bodies to practice medicine, the CPSO data contains both the universe of physicians who have ever worked in Ontario and the universe of Ontario physicians who have incorporated. The CPSM data contains the universe of individuals who are currently practicing in Manitoba and the universe of currently practicing physicians who have incorporated, as of 2025.

Second, for cross-provincial intensive margin outcomes, we use the National Physician Database Historical Utilization files that are publicly available through the Canadian Institute for Health Information (Canadian Institute for Health Information, 2021). These data consist of historical measures of services rendered and total payments to physicians in each province and territory by specialty. Data exists from 1990 to 2020 and we exclude territories.

Third, for cross-provincial extensive margin outcomes, we use the Supply, Distribution and Migration of Physicians in Canada. These data are publicly available through the Canadian Institute for Health Information (Canadian Institute for Health Information, 2020) and consist of historical measures of the annual number of physicians in each province and territory by specialty and by other characteristics. These include age of physician, graduation location, and others. We include data from 1970 to 2020 and we exclude territories.

### 3.2 Econometric Strategy

To assess the impact of the ability to incorporate on physician supply, we use staggered Callaway-Sant’Anna differences-in-differences estimation (Callaway and Sant’Anna, 2021). We compare treatment provinces that legislate physician incorporation and those that do not before and after the policy change occurs. We use year of legislation rather than regulation as treatment because it is often not clear for each province which year regulation occurred. We examine extensive margin outcomes up to 23 years and intensive margin outcomes up to 20 years after incorporation is allowed. In addition to the overall long-run effect, we provide two additional DiD estimates at 5 years (the “short-run” effect) and at 15 years of observation after incorporation is allowed (the “medium-run” effect).

We define treatment groups *g* ∈ {1975, 1981, 1988, 1990, 1996, 1999, 2000, 2001, 2005} based on the year each province first legislated incorporation. For each treatment group *g* and time period *t*, we estimate the group-time average treatment effect:

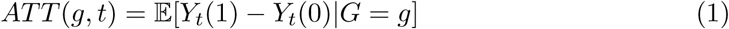

where *Y_t_*(1) is the potential outcome under treatment, *Y_t_*(0) is the potential outcome under control, and *G* = *g* indicates provinces first treated in period *g*.

The CS estimator uses variation in treatment timing to identify these effects, comparing treated units to appropriate control groups (never-treated units when available, or not-yet-treated units otherwise). We then aggregate the group-time effects to estimate:

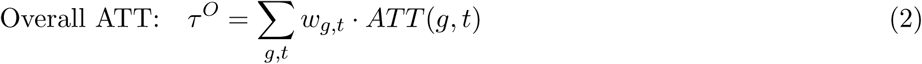

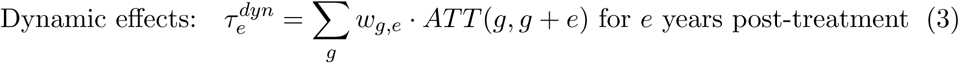

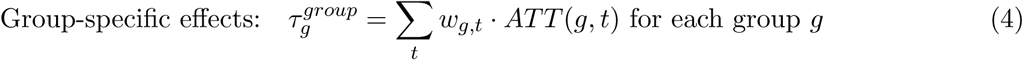

where *w_g,t_* are weights based on group sizes and time periods. This approach avoids the negative weighting problems that can arise in standard two-way fixed effects estimation with staggered treatment timing. There is a small clusters problem with only ten provinces (Cameron and Miller, 2015), so *ɛ* is a wild bootstrapped standard error.

For a DiD design to identify causal effects, two key assumptions must hold: (i) no other interventions occur at the time of treatment; and, (ii) treatment and control groups would have followed parallel trends in the absence of treatment. We assess the latter assumption using event-study analyses, testing for pre-trends with up to twenty years of placebo leads in our extensive margin specifications and eight years in our intensive margin specifications. In addition, we examine heterogeneity in treatment effects by physician age, years since graduation, and specialty, recognizing that the impact of incorporation is likely to differ by career stage and income level.

### 3.3 Outcomes

We first examine prevalence of incorporation in our CPSO and CPSM data. In our CPSO data, we describe the prevalence of incorporation in both provinces by proportion of a specialty who have incorporated and the proportion incorporated by year of graduation. We can also provide information on prevalence by number of physicians incorporated by year and years from graduation to incorporation. Our CPSM data only allows us to provide estimates of prevalence of incorporation.

For intensive margin outcomes in our DiD regressions, we are interested in the effect of allowing incorporation on the per capita number of services and the per capita payments provided in each province in each year. We examine outcomes by aggregated physician types. These include: 1) total physicians; 2) medical specialists; 3) surgical specialists; and, 4) family physicians. These categories are inherently interesting but because surgical specialists and medical specialists are better compensated than family physicians, they should exhibit larger labour supply responses. We examine how allowing incorporation affects types of aggregate services provided by the physician. These categories include: 1) number/payments for services; 2) number/payments for procedures; and, 3) number/payments for consultations and visits.

For extensive margin outcomes in our DiD regressions, we are interested in the effect of allowing incorporation on the supply of aggregate groups of physicians. These include: 1) total physicians; 2) medical specialists; 3) surgical specialists; and, 4) family physicians. We then examine how the supply of all physicians changes by age groups of physicians. We focus on physicians grouped into ages 30-39, 40-49, 50-59, 60-64, 65-69, and 70-74. Subsequently, we examine number of physicians by groups of physicians binned by year since graduation. These include physicians who are 6-10, 11-15, 16-20, 21-25, 26-30, and 31-35 years since graduation. Next, we assess whether the age structure of the physician labour force changes. These outcomes are the average age, median age, and median years from graduation of physicians within a province. Finally, to assess whether these policies incentivized physicians to move to a province, we also examine the number of physicians who have moved abroad, the number of physicians who have returned from abroad, and the net interprovincial migration of physicians.

We transform most outcomes into a logarithmic value to estimate an elasticity. In all cases, we convert the number of physicians or number/cost of services to a per 100,000 population measure using annual provincial population estimates. The exception to this is the foreign and domestic movement of physicians, which are often negative or zero, so cannot be log-transformed. In these cases, we estimate effects on the per capita outcome. Finally, we assess whether age structure of the physician labour force changes using the direct outcome.

## 4 Results

### 4.1 Adoption of Professional Incorporation in Ontario and Manitoba

We first present descriptive evidence on prevalence of incorporation. While we cannot observe prevalence of incorporation in our cross-provincial data, our Ontario CPSO and Manitoba CPSM data provides us with some confidence that when a province allows it, physician incorporation is widespread. Widespread uptake demonstrates that physicians view professional incorporation as in their interest.

For all physicians in our CPSO data, 40% have incorporated. Of individuals who are actively practicing in 2024 in Ontario, the number of physicians who incorporate rises to 68%. For individuals actively practicing in Manitoba as of 2025, 61% of all physicians are incorporated. Figure 2 demonstrates some descriptive statistics across several dimensions for our CPSO data. Incorporation only begins in 2003 after the policy is allowed. The peak number of physicians who incorporate occurs in 2020 when 3,000 did so. The cohort of physicians who are most likely to incorporate graduated in 1990. 60% of this cohort incorporated, while nearly 40% of the physicians who graduated from 2000 to 2015 incorporated.

**Figure 2:**
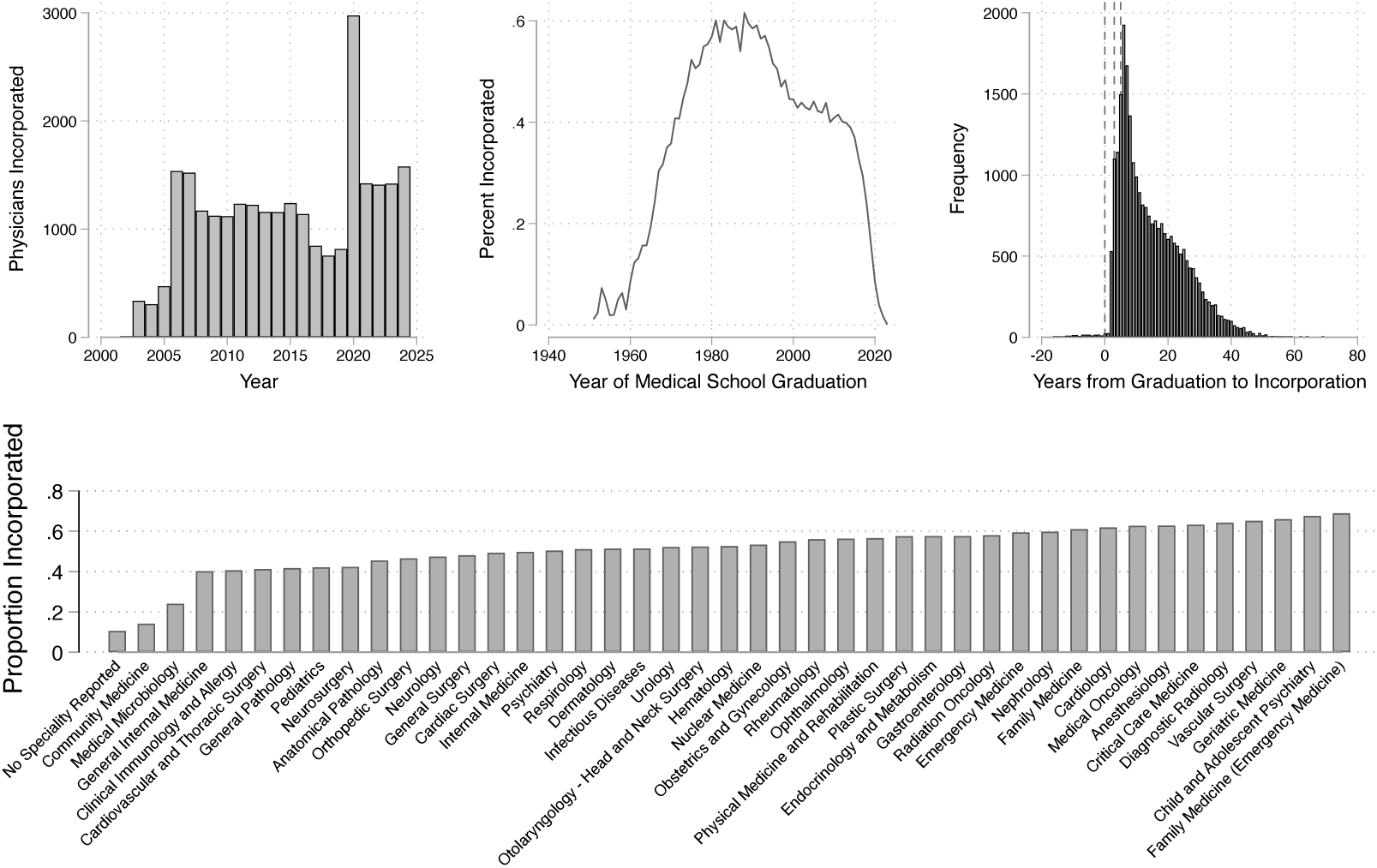
Descriptive statistics using College of Physicians and Surgeons of Ontario data on physician incorporation. Distribution of year of incorporation, proportion of individuals incorporated by graduation year, and years from graduation to incorporation in top panel. Proportion of physicians incorporating by specialty in bottom panel.

Most physicians who incorporate do so within 10 years of graduating medical school. However, there are substantial numbers of physicians who graduated before 1980 and who are at least 30 years out from graduating medical school who also incorporate. By specialty, the highest proportion of physicians that incorporate are family medicine practitioners who have also trained in emergency medicine. The lowest proportion of physicians who incorporate have no specialty reported. This is a historical artifact of a period when it was not necessary to complete a full residency to practice medicine in Canada.

### 4.2 Cross-provincial descriptive statistics

Table 2 shows summary statistics for our cross-provincial data on services and payments to physicians. For Canada, the average cost per service is 49.019 CAD, and there are 687,038.755 services provided per 100,000 population. The average price per procedure is 67.566 and there are 159,907.307 procedures per 100,000 population. The cost per consultation or visit is 44.061, and the average number of these consultations/visits is 527,131.405 per 100,000 population.

**Table 2:** Medical service supply statistics by province.

|  | Alberta | British Columbia | Manitoba | New Brunswick | Newfoundland and Labrador | Nova Scotia | Ontario | Prince Edward Island | Quebec | Saskatchewan | Total |
| --- | --- | --- | --- | --- | --- | --- | --- | --- | --- | --- | --- |
|  | **Total Services** |  |  |  |  |  |  |  |  |  |  |
| N | 26<br>(10.0%) | 26<br>(10.0%) | 26<br>(10.0%) | 26<br>(10.0%) | 26<br>(10.0%) | 26<br>(10.0%) | 26<br>(10.0%) | 26<br>(10.0%) | 26<br>(10.0%) | 26<br>(10.0%) | 260<br>(100.0%) |
| Cost per service | 61.037<br>(17.792) | 54.480<br>(10.837) | 45.943<br>(13.049) | 49.252<br>(11.950) | 40.492<br>(11.234) | 44.967<br>(8.973) | 46.931<br>(12.573) | 43.735<br>(16.053) | 56.395<br>(23.802) | 46.356<br>(13.111) | 49.021<br>(15.479) |
| Payments | 1,773,581,049<br>(892,047,056) | 1,884,218,714<br>(627,204,657) | 442,913,433<br>(198,194,189) | 233,426,546<br>(56,276,503) | 147,988,448<br>(48,933,071) | 251,909,302<br>(37,201,397) | 4,865,987,739<br>(1,176,260,319) | 31,261,477<br>(7,979,325) | 2,814,278,402<br>(1,243,055,509) | 357,802,164<br>(108,873,305) | 1,294,917,372<br>(1,631,224,683) |
| Services per 100K | 742,786<br>(68,869) | 752,252<br>(41,859) | 743,251<br>(80,900) | 630,534<br>(12,239) | 686,696<br>(54,232) | 607,011<br>(73,552) | 811,178<br>(93,382) | 522,790<br>(101,724) | 631,245<br>(33,055) | 723,694<br>(48,504) | 687,039<br>(103,274) |
| Total services | 27,283,340<br>(6,601,977) | 33,706,917<br>(4,797,781) | 9,214,123<br>(1,654,909) | 4,745,521<br>(91,048) | 3,603,728<br>(268,067) | 5,688,036<br>(628,427) | 104,502,738<br>(4,205,175) | 749,010<br>(113,009) | 49,574,276<br>(1,875,397) | 7,681,720<br>(377,010) | 24,954,232<br>(30,903,957) |
|  | **Total Procedures** |  |  |  |  |  |  |  |  |  |  |
| Cost per service | 80.121<br>(19.302) | 63.092<br>(10.310) | 65.876<br>(13.402) | 61.078<br>(8.872) | 52.476<br>(12.843) | 58.982<br>(9.231) | 55.300<br>(13.812) | 58.217<br>(11.681) | 76.745<br>(21.394) | 58.760<br>(11.763) | 63.098<br>(14.774) |
| Payments | 1,088,354,778<br>(584,204,015) | 1,060,156,443<br>(315,727,061) | 271,351,771<br>(144,902,166) | 131,164,958<br>(32,108,782) | 83,216,203<br>(31,914,960) | 144,286,847<br>(29,783,124) | 2,315,047,936<br>(721,076,276) | 19,362,472<br>(5,387,326) | 1,810,853,601<br>(888,337,393) | 205,186,567<br>(68,743,249) | 728,918,182<br>(1,059,298,391) |
| Services per 100K | 394,314<br>(47,175) | 429,551<br>(30,401) | 414,129<br>(63,016) | 341,859<br>(10,182) | 390,505<br>(49,062) | 353,545<br>(49,535) | 484,059<br>(59,801) | 320,607<br>(54,305) | 377,542<br>(31,920) | 392,091<br>(37,057) | 390,991<br>(54,822) |
| Total services | 13,330,321<br>(3,655,498) | 18,297,610<br>(2,629,009) | 5,427,529<br>(1,009,788) | 2,570,318<br>(86,246) | 2,056,470<br>(263,808) | 3,261,538<br>(407,800) | 49,550,798<br>(2,840,352) | 462,360<br>(71,043) | 23,586,211<br>(1,487,170) | 3,490,939<br>(289,441) | 13,646,177<br>(20,499,018) |
|  | **Consultations and Visits** |  |  |  |  |  |  |  |  |  |  |
| Cost per service | 42.731<br>(11.202) | 45.453<br>(6.309) | 29.176<br>(6.463) | 33.663<br>(7.090) | 28.847<br>(6.273) | 31.751<br>(5.889) | 34.425<br>(6.518) | 28.820<br>(11.157) | 39.594<br>(10.544) | 34.727<br>(8.624) | 34.229<br>(8.732) |
| Payments | 685,226,522<br>(316,762,981) | 823,178,946<br>(321,391,513) | 171,561,661<br>(77,720,873) | 101,191,588<br>(23,222,064) | 64,772,246<br>(20,236,949) | 107,622,454<br>(18,300,510) | 2,550,939,803<br>(586,764,265) | 11,898,957<br>(3,516,662) | 992,716,796<br>(379,417,332) | 152,615,598<br>(43,762,111) | 606,763,539<br>(654,099,537) |
| Services per 100K | 346,517<br>(31,021) | 322,700<br>(21,328) | 329,122<br>(29,850) | 288,675<br>(8,644) | 296,191<br>(15,779) | 253,466<br>(25,703) | 327,118<br>(39,722) | 202,184<br>(56,722) | 253,703<br>(15,617) | 319,755<br>(21,847) | 296,047<br>(48,452) |
| Total services | 13,953,019<br>(3,057,983) | 15,409,307<br>(2,030,105) | 3,786,593<br>(662,777) | 2,172,153<br>(39,869) | 1,547,258<br>(133,358) | 2,426,497<br>(260,074) | 74,951,940<br>(2,053,707) | 286,649<br>(54,957) | 25,166,482<br>(700,503) | 4,232,410<br>(183,634) | 17,725,980<br>(23,613,720) |

Table 3 shows summary statistics for our cross-provincial extensive margin data on number of physicians. The number of physicians per 100,000 population across Canada is 172.689. The average age of a physician over the period of observation is 46.998, and the median years from graduation is 17.825. On average, 33.1 physicians move abroad per year, whereas 23.9 return from abroad.

**Table 3:** Physicians supply statistics by province and type.

|  | All Physicians |  |  |  |  |  |  |  |  |  |  |
| --- | --- | --- | --- | --- | --- | --- | --- | --- | --- | --- | --- |
|  | Alberta | British Columbia | Manitoba | New Brunswick | Newfoundland and Labrador | Nova Scotia | Ontario | Prince Edward Island | Quebec | Saskatchewan | Total |
| N | 51<br>(10.0%) | 51<br>(10.0%) | 51<br>(10.0%) | 51<br>(10.0%) | 51<br>(10.0%) | 51<br>(10.0%) | 51<br>(10.0%) | 51<br>(10.0%) | 51<br>(10.0%) | 51<br>(10.0%) | 510<br>(100.0%) |
| Physician-to-100,000 pop. ratio | 171.747<br>(40.884) | 194.742<br>(26.775) | 172.669<br>(24.644) | 153.438<br>(48.382) | 170.878<br>(52.088) | 194.293<br>(45.758) | 180.103<br>(25.780) | 140.390<br>(30.984) | 196.372<br>(38.560) | 152.257<br>(27.183) | 172.689<br>(41.422) |
| Number of physicians | 5342.157<br>(2760.377) | 7365.529<br>(2667.249) | 1999.882<br>(463.057) | 1137.216<br>(399.123) | 926.412<br>(258.073) | 1778.882<br>(495.685) | 20373.745<br>(6533.317) | 190.471<br>(57.789) | 14459.745<br>(4096.898) | 1569.157<br>(391.328) | 5514.320<br>(6994.521) |
| Number male | 3783.353<br>(1466.266) | 5352.529<br>(1300.059) | 1485.627<br>(193.126) | 824.882<br>(181.728) | 676.039<br>(121.189) | 1275.314<br>(208.932) | 14567.098<br>(2899.247) | 149.412<br>(30.738) | 9813.451<br>(1301.138) | 1185.569<br>(169.295) | 3911.327<br>(4681.151) |
| Number female | 1543.961<br>(1314.070) | 1986.647<br>(1429.235) | 500.451<br>(297.636) | 300.588<br>(236.612) | 241.078<br>(153.968) | 493.431<br>(311.354) | 5701.627<br>(3847.470) | 39.196<br>(30.073) | 4576.451<br>(3207.670) | 367.804<br>(245.090) | 1575.124<br>(2539.569) |
| Average age | 45.869<br>(1.440) | 47.237<br>(2.130) | 47.263<br>(1.708) | 46.765<br>(1.650) | 44.973<br>(2.659) | 47.404<br>(2.201) | 47.749<br>(2.128) | 48.824<br>(2.170) | 46.763<br>(2.261) | 47.139<br>(1.494) | 46.998<br>(2.230) |
| Median age | 44.569<br>(1.982) | 46.333<br>(2.696) | 46.039<br>(2.308) | 45.549<br>(2.016) | 43.667<br>(3.029) | 46.333<br>(2.805) | 46.608<br>(2.616) | 48.167<br>(2.712) | 45.431<br>(3.022) | 46.020<br>(1.435) | 45.872<br>(2.740) |
| Number in rural areas | 487.941<br>(185.959) | 496.745<br>(192.548) | 264.510<br>(83.427) | 201.196<br>(74.327) | 247.235<br>(83.379) | 361.294<br>(122.558) | 962.039<br>(337.651) | 27.216<br>(8.663) | 1390.294<br>(532.480) | 235.882<br>(74.487) | 467.435<br>(449.451) |
| Number in urban areas | 4679.529<br>(2833.525) | 6590.608<br>(2993.177) | 1629.765<br>(616.889) | 886.176<br>(415.695) | 638.647<br>(265.510) | 1335.980<br>(547.860) | 18522.725<br>(7907.009) | 154.667<br>(68.344) | 12447.765<br>(4895.679) | 1245.922<br>(516.076) | 4813.178<br>(6651.333) |
| Place of MD graduation: Canada | 3559.627<br>(1927.170) | 5035.294<br>(2077.264) | 1251.824<br>(376.913) | 819.510<br>(344.713) | 496.627<br>(218.585) | 1219.569<br>(407.892) | 14333.235<br>(5298.504) | 144.667<br>(46.458) | 12324.745<br>(4125.485) | 713.706<br>(225.842) | 3989.880<br>(5420.819) |
| Place of MD graduation: Foreign | 1726.373<br>(907.774) | 2229.020<br>(730.989) | 702.784<br>(134.190) | 288.392<br>(92.438) | 417.353<br>(64.414) | 520.569<br>(143.819) | 5663.922<br>(1810.739) | 34.882<br>(8.342) | 1726.725<br>(321.668) | 820.294<br>(203.096) | 1413.031<br>(1718.626) |
| Median years since graduation | 16.784<br>(3.221) | 18.275<br>(4.181) | 17.863<br>(4.030) | 17.510<br>(3.603) | 16.284<br>(4.181) | 17.980<br>(4.379) | 18.451<br>(4.220) | 18.578<br>(5.285) | 18.294<br>(4.601) | 18.225<br>(3.180) | 17.825<br>(4.158) |
| Returned from abroad | 26.080<br>(10.202) | 37.740<br>(12.584) | 10.420<br>(5.230) | 3.500<br>(2.188) | 3.420<br>(1.980) | 9.360<br>(4.014) | 90.300<br>(30.323) | 0.800<br>(0.990) | 49.360<br>(19.067) | 8.100<br>(4.344) | 23.908<br>(29.780) |
| Moved abroad | 34.700<br>(19.257) | 43.340<br>(22.014) | 17.220<br>(11.995) | 5.820<br>(3.832) | 5.860<br>(4.352) | 14.600<br>(10.833) | 130.840<br>(68.477) | 0.740<br>(0.965) | 61.280<br>(33.291) | 16.900<br>(10.822) | 33.130<br>(45.619) |
| Net migration within Canada | 12.902<br>(45.195) | 71.392<br>(56.033) | -24.706<br>(16.438) | -1.784<br>(10.616) | -23.431<br>(15.599) | -6.353<br>(14.657) | 23.529<br>(56.422) | 1.275<br>(4.299) | -21.118<br>(41.648) | -31.176<br>(20.668) | 0.053<br>(44.319) |

### 4.3 Cross-Provincial Difference-in-Differences Results

Table 4 demonstrates overall DiD effects for services after incorporation is allowed for intensive margin outcomes. Across most physician types and services provided, there is little effect of allowing physician incorporation. However, surgical specialists reduce their total services by 13.3% and procedures by 21.2%. Figure 3 shows event study results by all physicians and physician subtype. Both our event analysis and short/medium run DiD estimates suggest that significant effects develop within five years and increase in magnitude over time.

**Figure 3:**
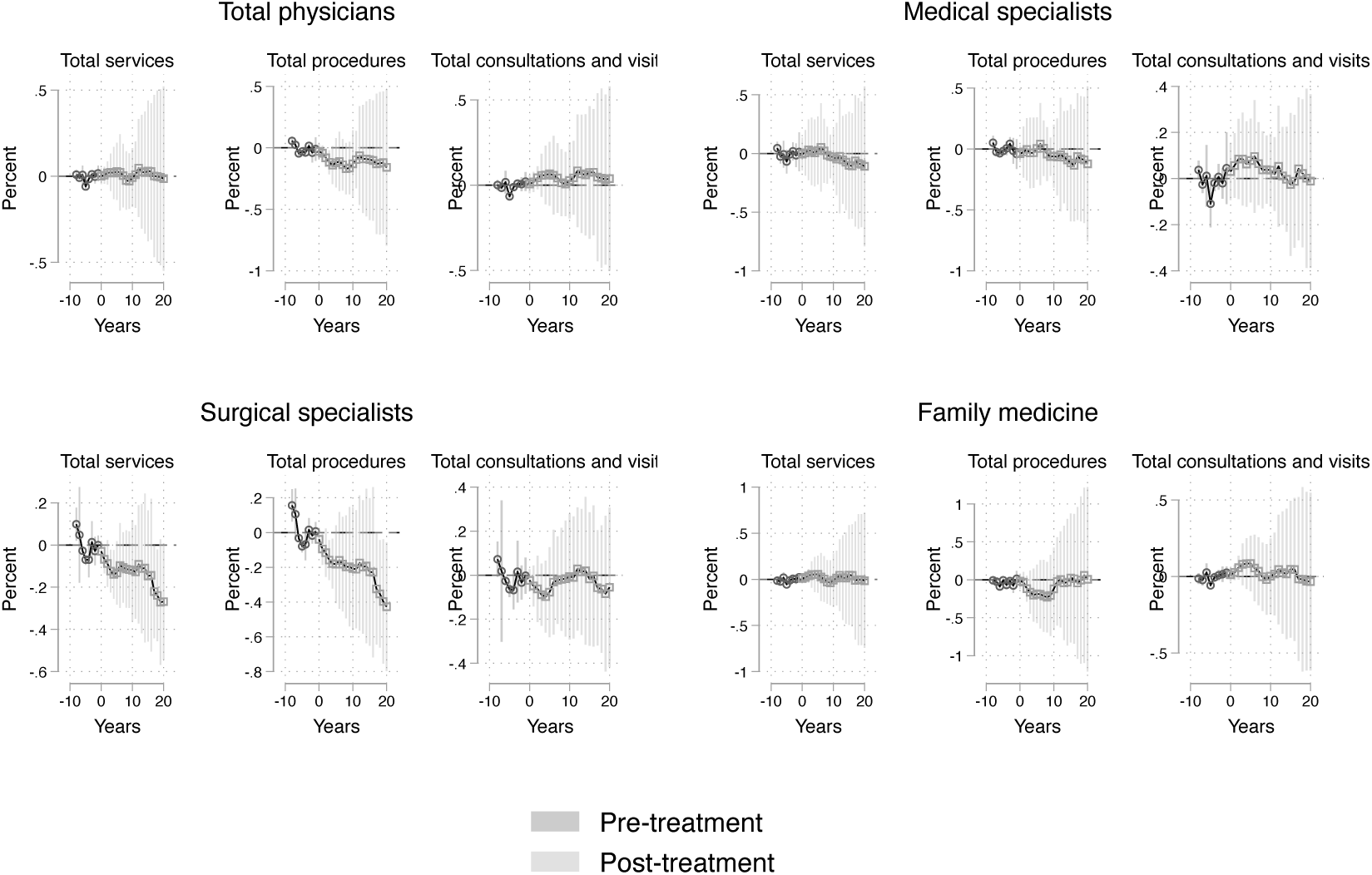
Event studies of services provided, by specialty type, when allowing incorporation at time zero.These compare provinces that allow incorporation to those that do not before and after allowing physician incorporation. The estimated effect is an elasticity of the number of services, procedures, and consultations/visits. Estimates are in black and 95% CI are displayed in grey.

**Table 4:**
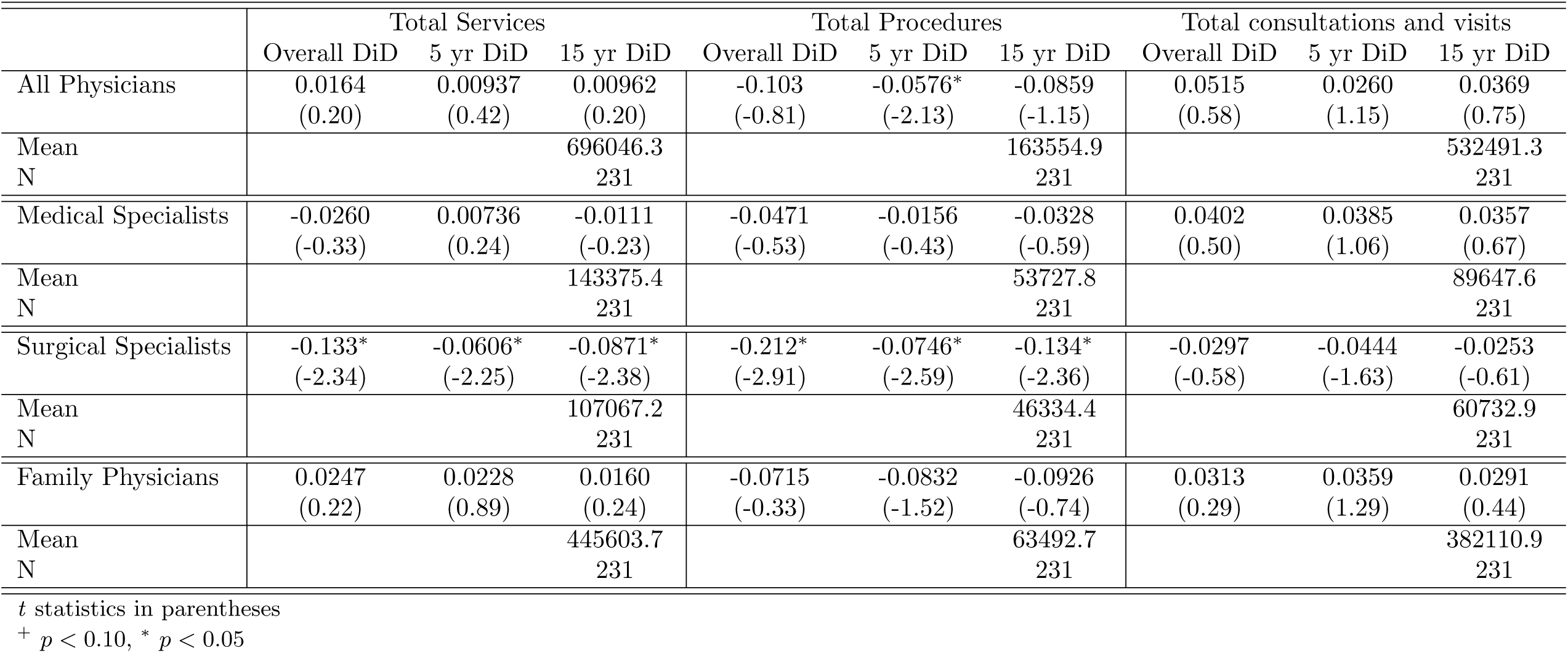
Overall DiD effects on service provision by type of physician.

Table 5 and Figure 4 demonstrates the effect of allowing incorporation on payments to physicians. Most results are not statistically significant and point estimates are small in magnitude. We find weak evidence that there are decreases in payments to surgical specialists at 5 years after incorporation is allowed, but this effect disappears in the overall and 15 year DiD estimates. We find little evidence of pre-trends in any of our event study graphs for intensive margin outcomes.

**Figure 4:**
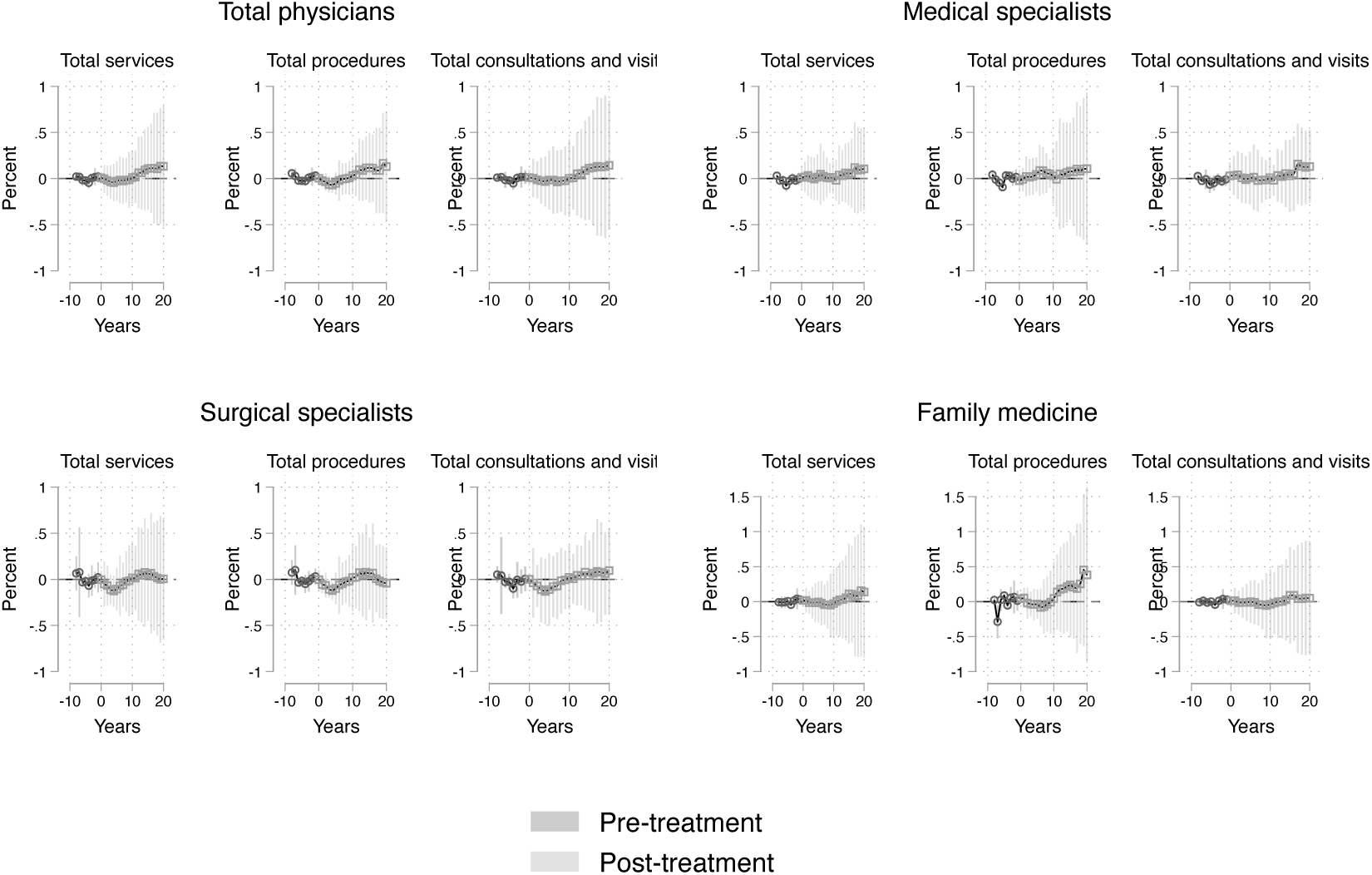
Event studies of payments provided, by specialty type, when allowing incorporation at time zero.These compare provinces that allow incorporation to those that do not before and after allowing physician incorporation. The estimated effect is an elasticity of the payments for services, procedures, and consultations/visits. Estimates are in black and 95% CI are displayed in grey.

**Table 5:**
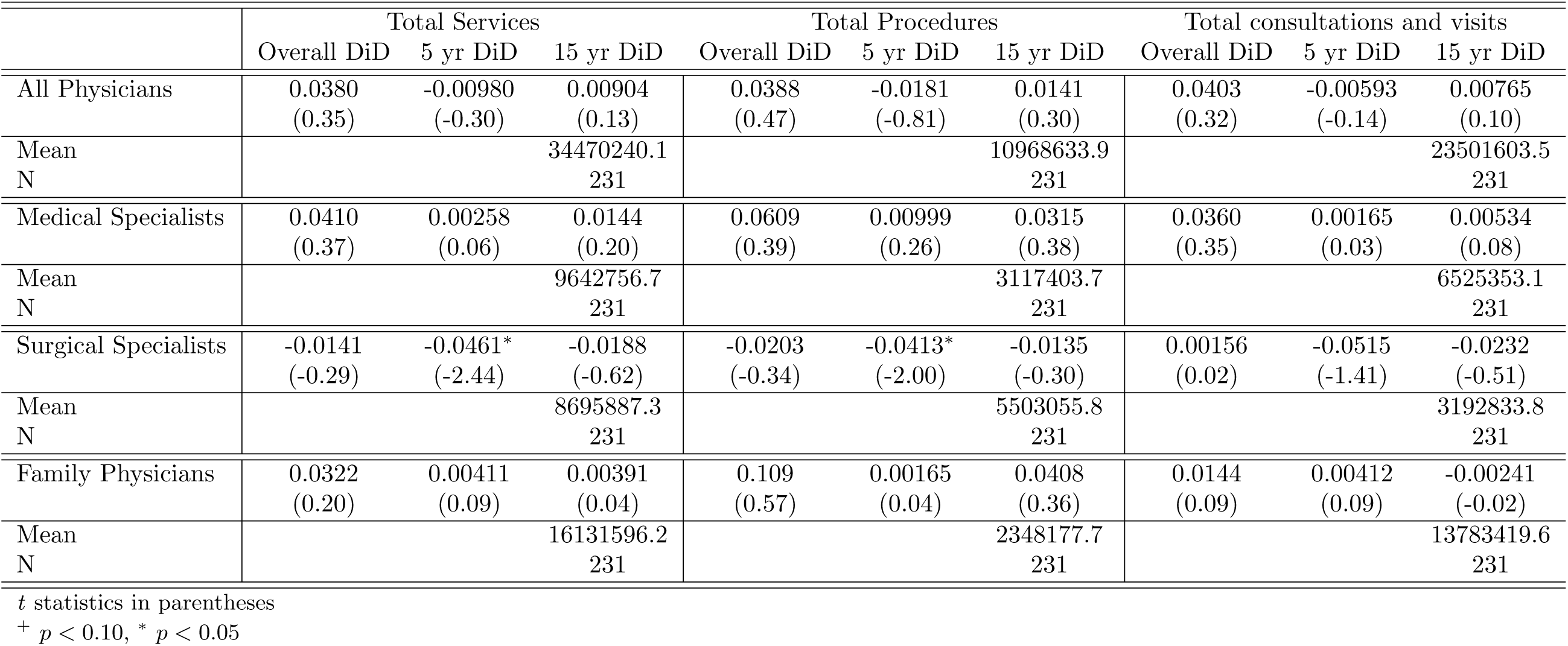
Overall DiD effects on payments by type of physician.

We next estimate extensive margin effects of incorporation. Our overall point estimates in Table 6 suggest that incorporation reduced all physicians, medical specialists, and surgical specialists by 6.8%, 7.9%, and 12.7%, respectively. Family physician extensive margin supply is not affected by allowing incorporation. Our short and medium run DiDs suggest these develop within five years and increase over time.

**Table 6:** Overall DiD effects on extensive supply of labour by type of physician. Coefficients are interpretable as elasticities.

|  | All Physicians |  |  | Medical Specialists |  |  | Surgical Specialists |  |  | Family Physicians |  |  |
| --- | --- | --- | --- | --- | --- | --- | --- | --- | --- | --- | --- | --- |
|  | Overall Effect | 5 yr DiD | 15 yr DiD | Overall Effect | 5 yr DiD | 15 yr DiD | Overall Effect | 5 yr DiD | 15 yr DiD | Overall Effect | 5 yr DiD | 15 yr DiD |
| ATT | -0.0682* | -0.0192* | -0.0353* | -0.0796* | -0.0227* | -0.0423* | -0.127* | -0.0249* | -0.0588* | -0.0466 | -0.0158<sup>+</sup> | -0.0255 |
|  | (-2.01) | (-2.81) | (-2.12) | (-2.93) | (-2.99) | (-2.90) | (-2.73) | (-2.43) | (-2.66) | (-0.92) | (-1.68) | (-1.01) |
| Mean |  |  | 153.8 |  |  | 45.54 |  |  | 23.35 |  |  | 84.75 |
| N |  |  | 350 |  |  | 350 |  |  | 350 |  |  | 350 |

*t* statistics in parentheses
<sup>+</sup> $p < 0.10$ , \* $p < 0.05$

Our events provide similar evidence, although they do not become individually significant until about 20 years after implementation. Figure 5 demonstrates event studies across four of the aggregate physician groups. These suggest that allowing incorporation is associated with declines in physician supply by 5 to 10% within five years after the policy is adopted. These occur for all physicians and surgical specialists. We find no evidence of pre-trends in these event studies.

**Figure 5:**
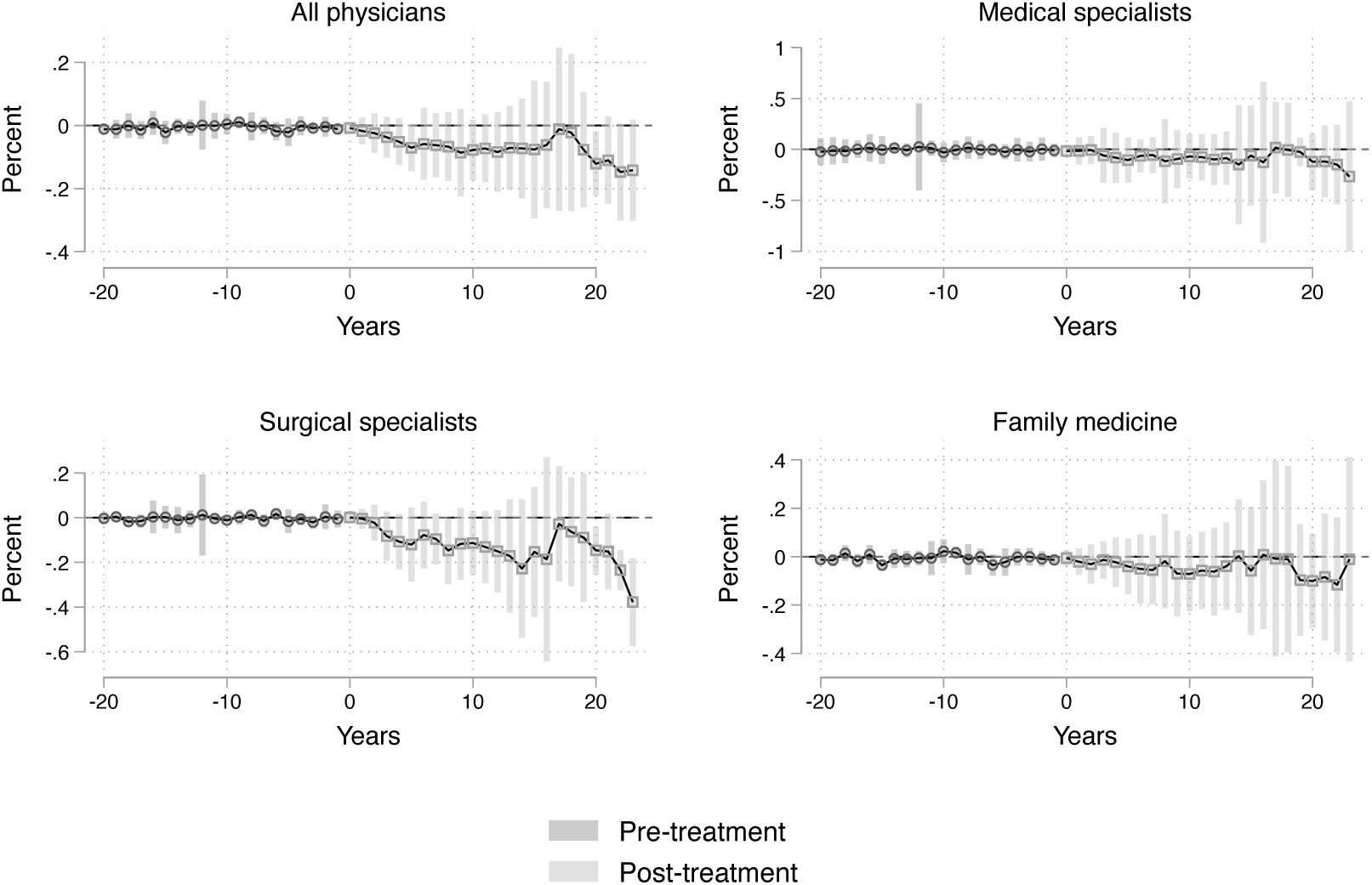
Event studies of extensive margin physician supply elasticities when allowing incorporation at time zero. These compare provinces that allow incorporation to those that do not before and after allowing physician incorporation. Estimates are in black and 95% CI are displayed in grey.

These results are primarily driven by reductions in older physicians and those later in their careers. Table 7 and 8 demonstrate our DiDs for age cohorts and cohort by year of graduation. We find estimated overall reductions in extensive margin physician supply in 40-49 and 60-64 age cohorts by 11.3 and 17.5%, respectively. We also find marginally significant reductions in supply of 65-69 and 70-74 year-olds by 17.2 and 20.1%, respectively. Cohorts further from graduation are also more affected by allowing incorporation. Those 31-35 years out from graduation reduce supply by 14.3%, although we find marginal reductions in cohorts that are 11-15 years from graduation as well.

**Table 7:**
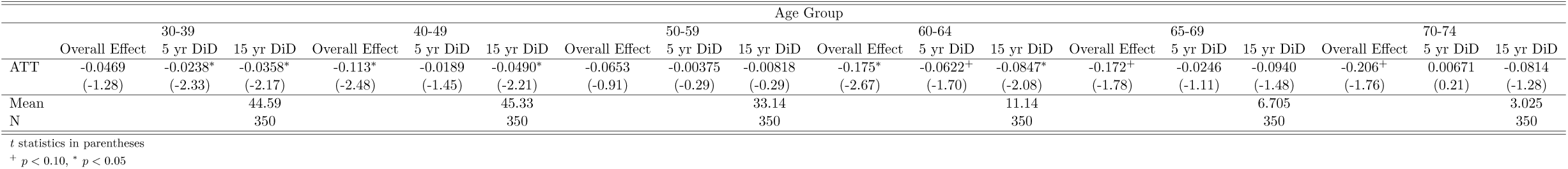
Overall DiD effects on extensive supply of labour by age group of physician. Coefficients are interpretable as elasticities.

**Table 8:**
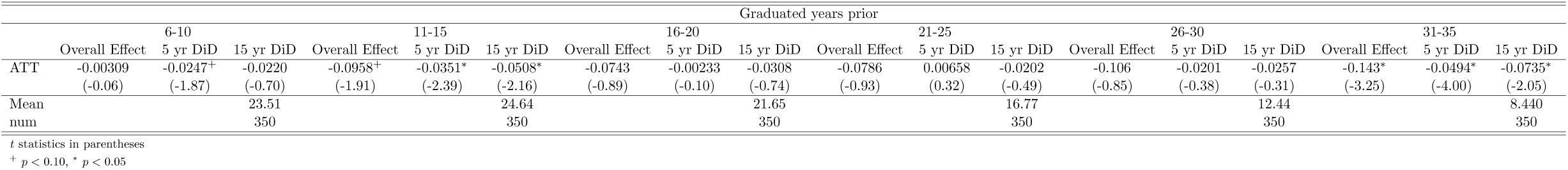
Overall DiD effects on extensive supply of labour by graduation group of physician. Coefficients are interpretable as elasticities.

Figure 6 demonstrates event studies for all physicians by age group. At 20 years after implementation, our events demonstrate significant reductions in physician extensive margin labour supply especially in cohorts aged 50 to 64. Figure 7 demonstrates effects by graduation cohort. Physicians who are 21 to 35 years out from graduating medical school demonstrate similar reductions 20 years after a province allows incorporation.

**Figure 6:**
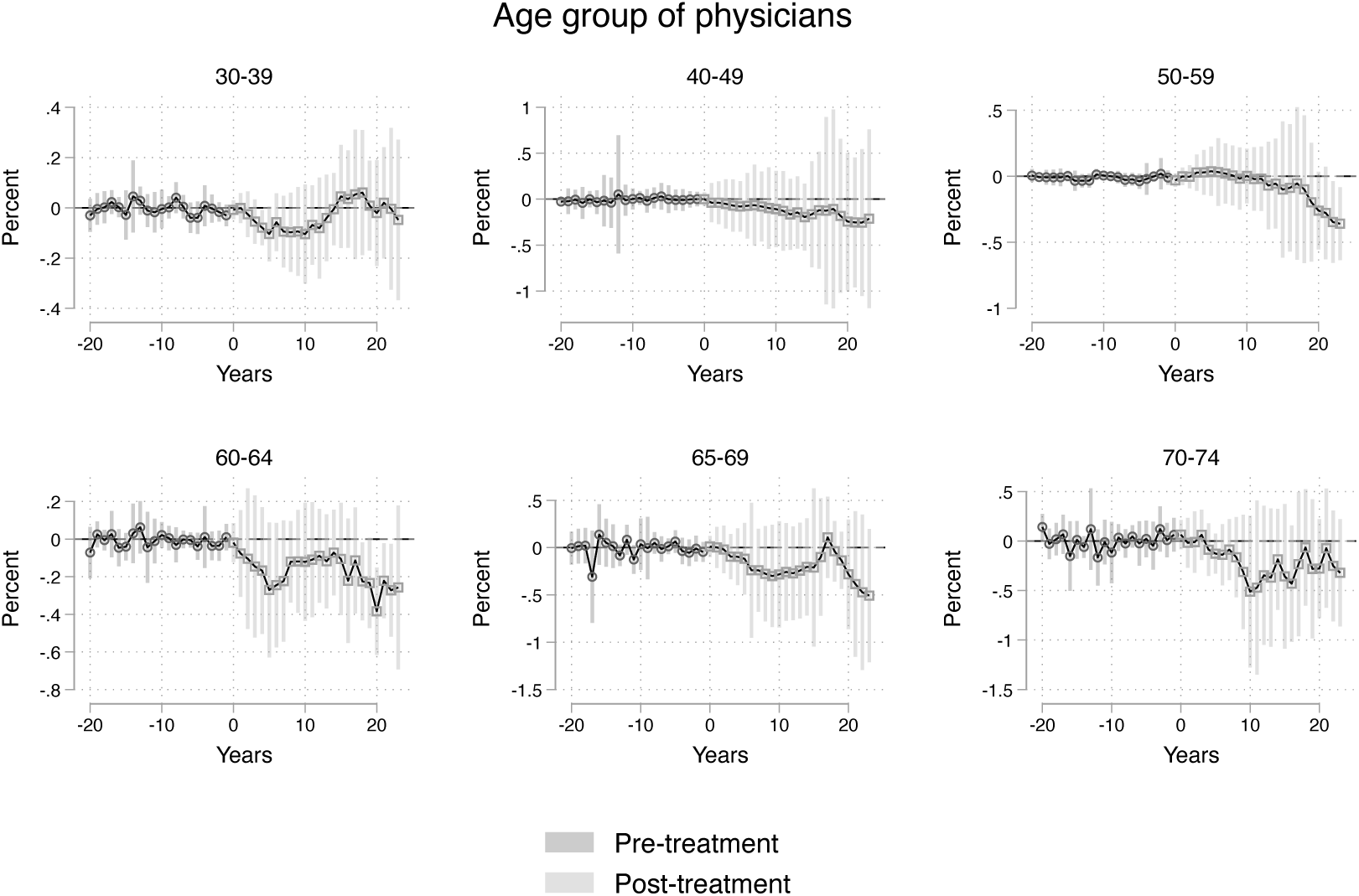
Event studies of extensive margin physician supply elasticities when allowing incorporation at time zero by age group of physicians These compare provinces that allow incorporation to those that do not before and after allowing physician incorporation. Estimates are in black and 95% CI are displayed in grey.

**Figure 7:**
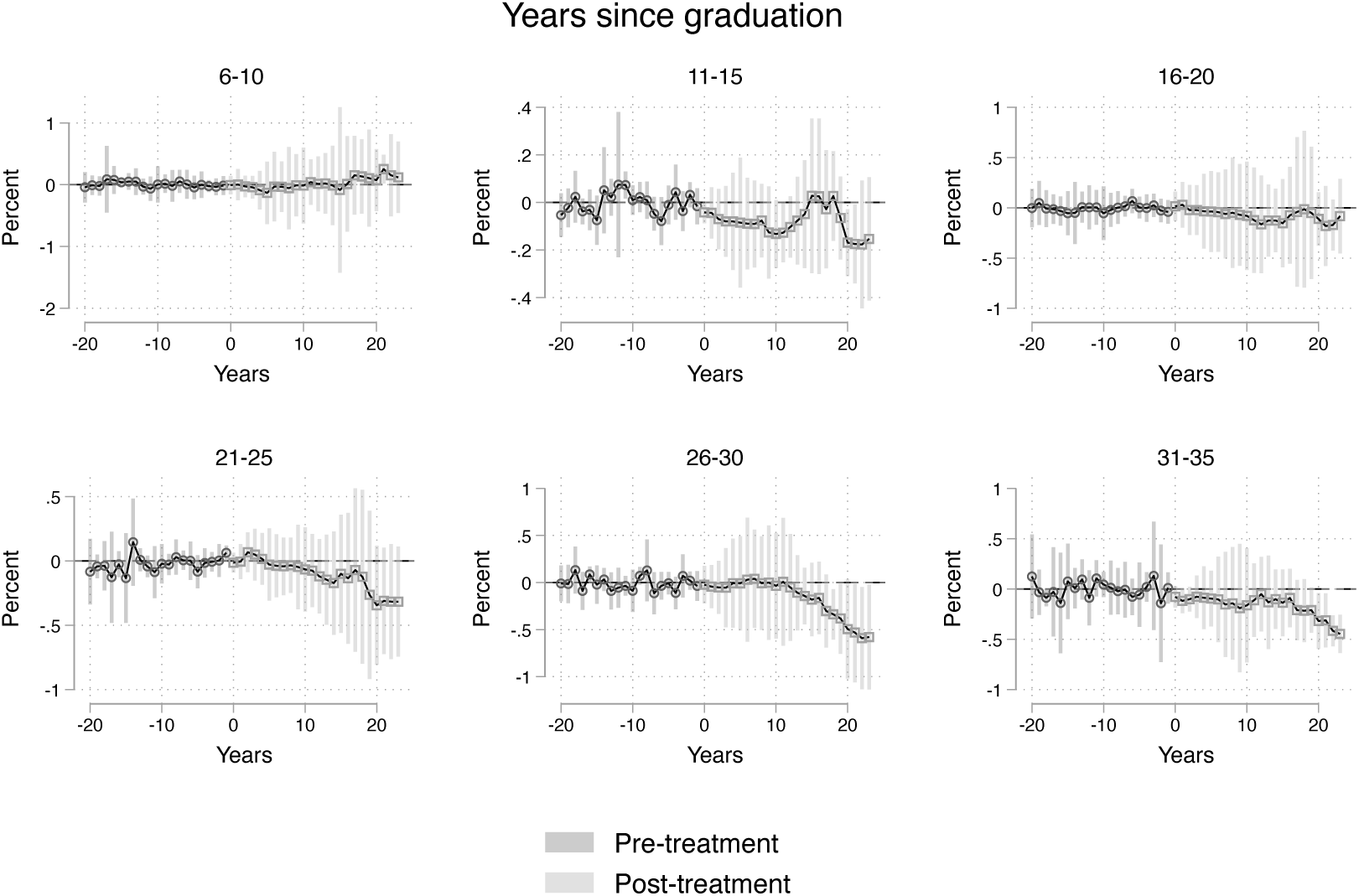
Event studies of extensive margin physician supply elasticities when allowing incorporation at time zero by years from graduation of physicians. These compare provinces that allow incorporation to those that do not before and after allowing physician incorporation. Estimates are in black and 95% CI are displayed in grey.

Reinforcing these results is evidence that the age structure of physicians changes in response to provinces allowing incorporation. Figure 8 and Table 9 demonstrate events and DiD estimates of how allowing incorporation causes changes in the average, and median age and median age since graduation. The overall effect is to reduce the average age of physicians by 0.747 years and the median age of physicians by 1.1 years. It also marginally reduces the median years since graduation of a physician practicing by 0.5 years. These effects are evident in statistically significant individual events approximately 20 years after policy implementation.

**Figure 8:**
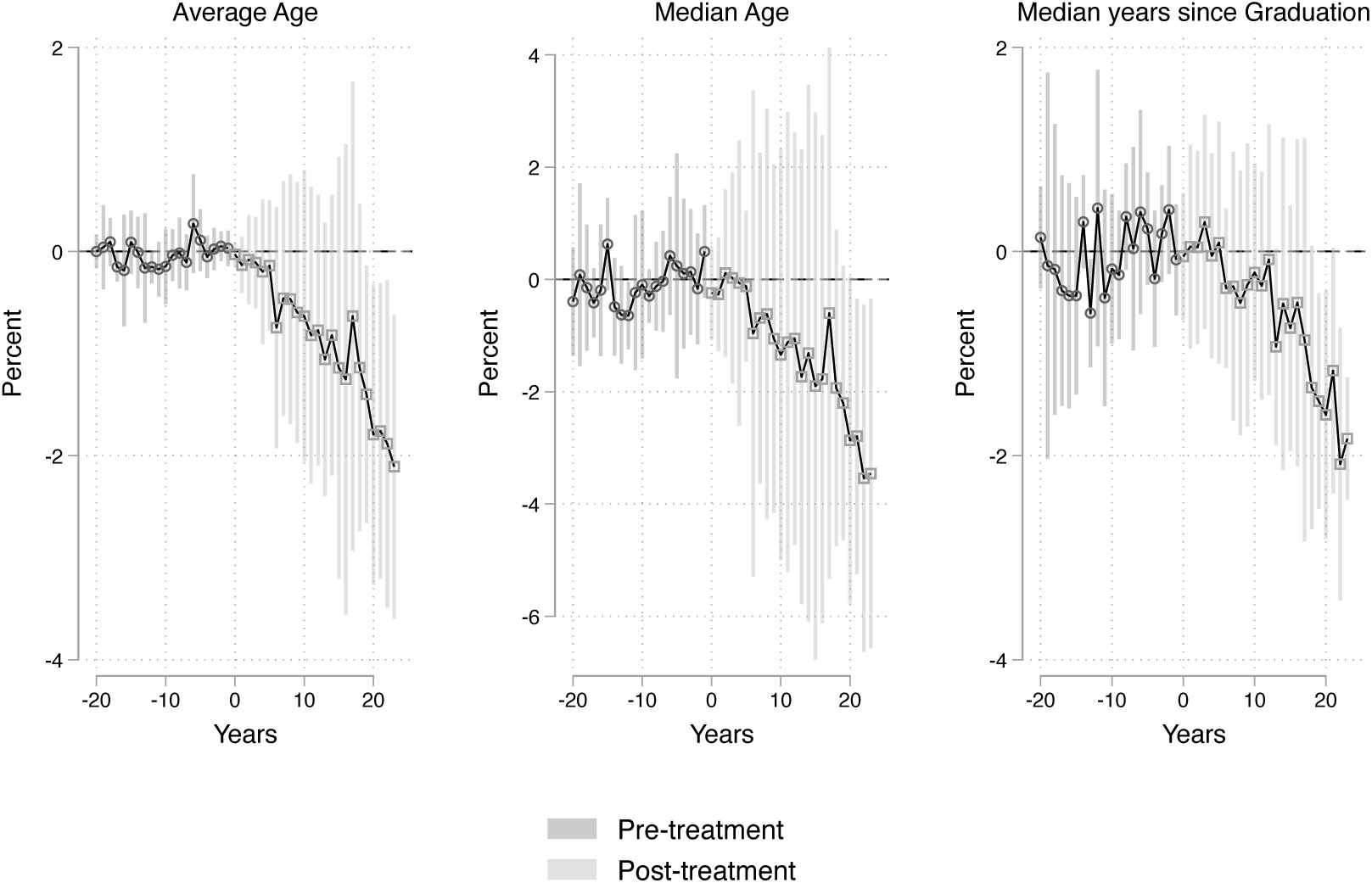
Event studies of extensive margin physician supply elasticities when allowing incorporation at time zero by age and graduation means and medians. These compare provinces that allow incorporation to those that do not before and after allowing physician incorporation. Estimates are in black and 95% CI are displayed in grey.

**Table 9:**
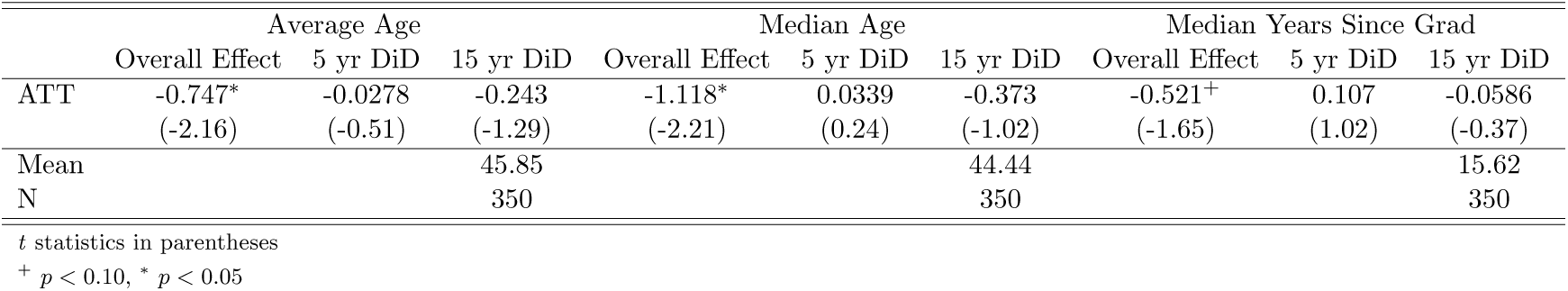
Overall DiD effects on age variables. Coefficients are interpretable as change in outcome.

Finally, we find modest changes for measures of physician migration caused by allowing incorporation. Table 10 documents a 0.688 physician per 100,000 population reduction in physicians moving abroad after allowing incorporation. There is a 1.142 physician per 100,000 population increase in the number of physicians immigrating into a province after allowing incorporation, although this is only statistically significant at the 10% level. Event studies, demonstrated in Figure 9, for these physician outcomes are noisy.

**Figure 9:**
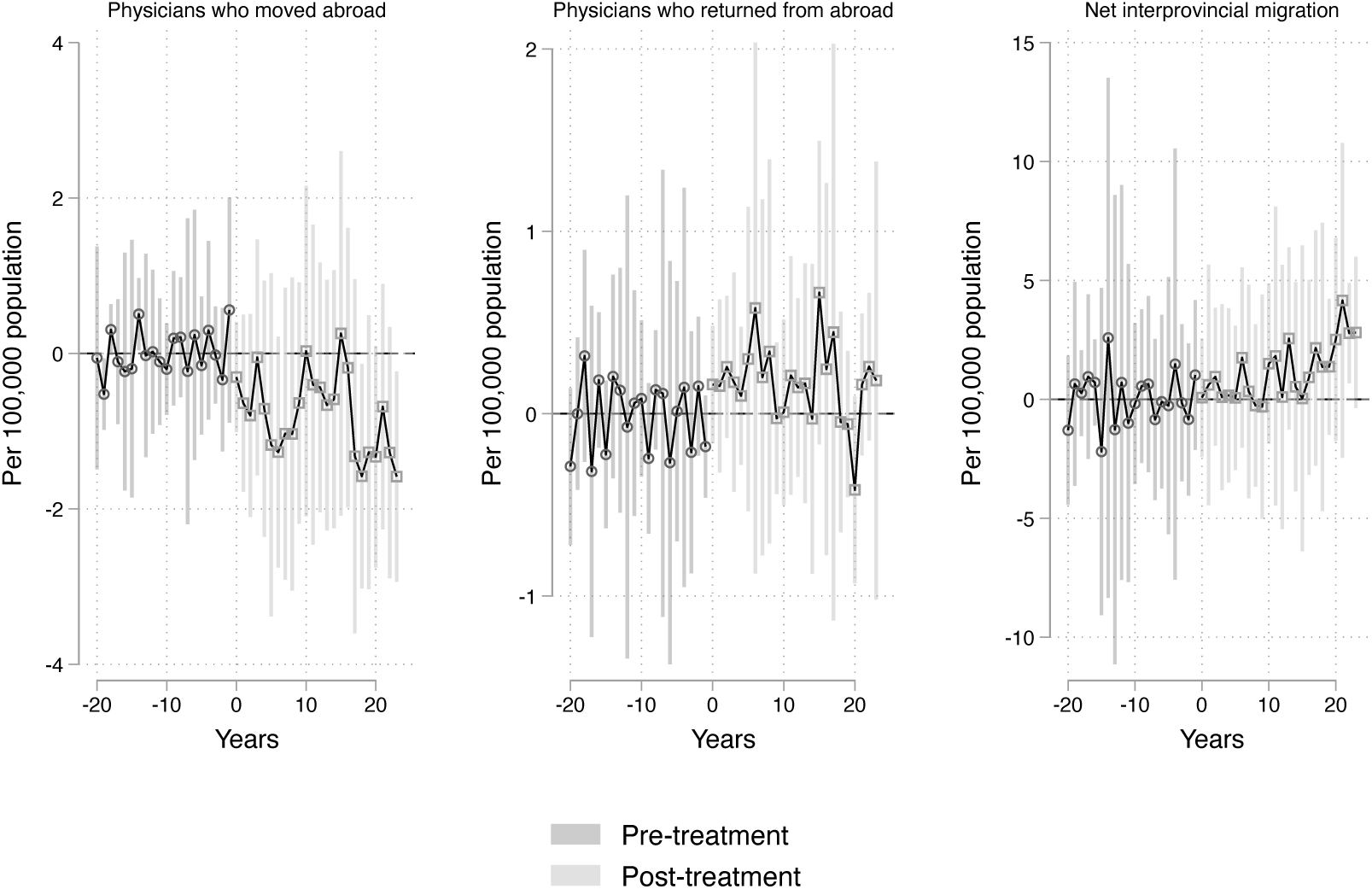
Event studies of extensive margin physician supply elasticities when allowing incorporation at time zero by migration patterns. These compare provinces that allow incorporation to those that do not before and after allowing physician incorporation. Estimates are in black and 95% CI are displayed in grey.

**Table 10:** Overall DiD effects on physician migration. Coefficients are interpretable as change in per 100k population.

|  | MDs who moved abroad |  |  | MDs who returned |  |  | Net interprovincial migration |  |  |
| --- | --- | --- | --- | --- | --- | --- | --- | --- | --- |
|  | Overall Effect | 5 yr DiD | 15 yr DiD | Overall Effect | 5 yr DiD | 15 yr DiD | Overall Effect | 5 yr DiD | 15 yr DiD |
| ATT | -0.688* | -0.211<sup>+</sup> | -0.337<sup>+</sup> | 0.183 | 0.0719<sup>+</sup> | 0.119<sup>+</sup> | 1.142<sup>+</sup> | 0.221 | 0.400 |
|  | (-2.09) | (-1.71) | (-1.76) | (1.52) | (1.89) | (1.67) | (1.89) | (1.27) | (1.52) |
| Mean |  |  | 1.500 |  |  | 0.877 |  |  | -0.597 |
| N |  |  | 350 |  |  | 350 |  |  | 350 |

*t* statistics in parentheses
<sup>+</sup> $p < 0.10$ , \* $p < 0.05$

## 5 Conclusion

This paper examines how professional incorporation affects physician labour supply in Canada. By allowing physicians to defer personal income taxation through corporate retention of earnings, incorporation substantially altered their financial incentives surrounding career and retirement choices. Evidence from Ontario and Manitoba shows that when policy permits, the majority of physicians incorporate.

Our cross-provincial difference-in-differences analysis reveals that incorporation reduces rather than increases labour supply. Intensive margin responses are limited overall but pronounced among surgical specialists, whose service provision declines significantly. Extensive margin effects are larger and persistent: total physician supply falls by nearly 7% in the long run, with reductions concentrated among older physicians and those later in their careers. The age profile of the workforce shifts accordingly, backing up the theory that this policy facilitated retirement.

These findings are consistent with economic theory emphasizing the tension between substitution and income effects of this particular policy. Policies that improve the effective return to work should, in principle, encourage greater effort. However, when policies primarily increase lifetime wealth, income effects can dominate. In the case of physician incorporation, enhanced retirement financing appears to have facilitated earlier workforce exit rather than additional labour supply.

Our empirical cross-provincial findings support theoretical predictions that income effects dominate substitution effects when tax deferral policies increase lifetime wealth, particularly for high earners near retirement thresholds. This is precisely what we observe: incorporation reduces rather than increases labour supply, with the largest effects among high-earning surgical specialists and physicians approaching retirement age.

Our findings have important implications for tax policy design beyond healthcare. The results demonstrate how policies intended to increase professional labour supply can backfire when they primarily increase lifetime wealth rather than current work incentives. This insight applies broadly to tax deferral policies across professions and jurisdictions.

The key mechanism is that tax deferral policies create wealth effects by improving retirement financing, which can dominate any substitution effects from higher effective returns to current work. This is particularly likely when: (1) policies target high earners who already have substantial retirement savings; (2) the tax benefits are large relative to current income; and, (3) the target population includes many individuals already considering retirement. Similar professional incorporation policies exist for lawyers, accountants, and other high-skilled professionals across many countries. Our results suggest these policies may systematically reduce rather than increase professional labour supply through facilitated early retirement, particularly as professional workforces age globally.

While we do not formally analyze welfare in this paper, the effect of allowing incorporation are likely negative. The 6.8% reduction in physician supply translates to approximately 3,749 fewer physicians nationwide which likely increased wait times and reduced healthcare access. Fiscal costs include both forgone tax revenue from early retirements and the direct costs of tax deferral benefits. The distributional effects may be particularly concerning: the policy transfers resources from general taxpayers to highearning professionals while reducing the supply of essential services. The benefits accrue to physicians who would likely have adequate retirement savings regardless, while the costs fall on patients who face reduced healthcare access.

Our findings raise important questions about the political economy of professional tax incentives. If these policies systematically reduce rather than increase professional labour supply, why do they persist and spread across jurisdictions? Several factors may explain this puzzle. Professional associations have strong incentives to lobby for tax benefits, while the costs are diffuse across taxpayers and patients. Policymakers may focus on the visible benefits (professional retention in the short run) while ignoring harderto-observe costs (earlier retirement in the long run). The complexity of tax deferral mechanisms may also obscure their true effects from both policymakers and the public. Understanding these political economy dynamics is crucial for designing more effective policies to address professional shortages.

Rather than policies that increase lifetime wealth and facilitate early retirement, effective interventions should target the specific constraints that limit professional labour supply. Direct retention bonuses tied to continued practice would create substitution effects without the problematic wealth effects of tax deferral. Structured career transition programs could address the discrete nature of retirement decisions by allowing gradual workforce exit. Improving practice conditions and reducing administrative burdens could extend careers without creating perverse financial incentives. The key insight is that effective policies should increase the marginal benefit of current work rather than the returns to retirement savings. These may be more expensive in the short-run but better for welfare in the long-run.

Several limitations suggest directions for future research. First, our focus on physicians provides a clean natural experiment but limits generalizability to other professions with different income levels, retirement patterns, and work arrangements. Studies of lawyer and accountant incorporation would test whether our findings extend to other high-skilled professionals. Second, we cannot observe the extensive margin of medical school applications or specialty choice. If incorporation affects career entry decisions, our estimates may understate the full labour supply effects. Future work combining our approach with medical education data could quantify these selection effects.

## Data Availability

All data produced in the present study are available upon reasonable request to the authors

## B Theoretical Model

### B.1 Modeling the Physician Labour-Leisure Decision

To understand our empirical findings, we develop a two-period model in which physicians choose labour supply in each period and make intertemporal consumption decisions. Professional incorporation affects their budget constraint by improving the effective return to deferred consumption, creating both income and substitution effects whose relative magnitudes determine the overall labour supply response.

#### B.1.1 Setup

Physicians live for two periods with heterogeneous preferences and lifecycle positions. In each period *t* = 1, 2, they choose labour supply *h_t_* ∈ [0, 1] and consumption *C_t_*. Period 1 represents prime working years; period 2 represents the pre-retirement period where physicians decide whether to continue working (*h*_2_ *>* 0) or retire early (*h*_2_ = 0). Physicians differ in their discount factor *δ*, with older physicians having lower *δ* (closer to retirement), and in their leisure preference *ϕ*. Preferences are additively separable and described by a Stone–Geary utility function (Blundell et al., 1993; Cesarini et al., 2017):

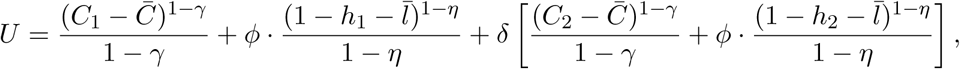

where *γ >* 0 governs intertemporal substitution, *η >* 0 governs the marginal utility of leisure, *ϕ >* 0 weights leisure preference relative to consumption, and *δ* ∈ (0, 1) is the time discount factor. The parameters ̄*C >* 0 and ̄*l* > 0 represent subsistence levels of consumption and leisure, respectively. Utility is only defined when *C_t_ > ̄C* and *h_t_ <* 1 − ̄*l*, ensuring agents meet basic needs before receiving marginal utility from additional consumption or leisure.

#### B.1.2 Budget Constraints and Incorporation

**Without incorporation**, all income is taxed immediately at a personal rate *τ_p_*:

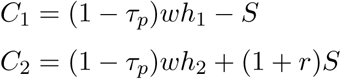

where savings *S* must be made from after-tax income, but accumulated savings are not taxed again in period 2.

**With incorporation**, only consumed income is taxed:

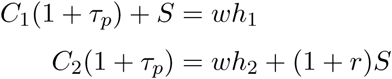

which can be rewritten as:

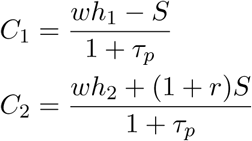

where savings *S* can be made from pre-tax income, and consumption is taxed when it occurs.

The key first difference from allowing incorporation is that it permits savings from pre-tax income, effectively increasing the amount available for investment. This creates a higher effective return to deferred consumption: without incorporation, each dollar of period 1 consumption sacrificed yields (1 + *r*) dollars in period 2, while with incorporation it yields (1 + *r*)*/*(1 − *τ_p_*) dollars in period 2 after taxes.

The second key difference is that incorporation makes it advantageous to reduce work effort in period 2 because the tax rate in each period depends on taxable income in that period. We assume two tax rates where:

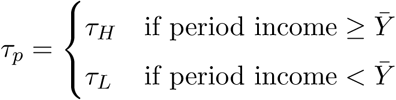

In period 1, the tax rate depends on *wh*_1_. In period 2, the tax rate depends on *wh*_2_ + (1 + *r*)*S* with incorporation (or *wh*_2_ without incorporation). Physicians can strategically choose *h*_2_ to potentially qualify for the lower rate *τ_L_* in period 2.

#### B.1.3 First-Order Conditions

We focus on the economically relevant comparison for physicians who typically earn high income in period 1 (*wh*_1_ ≥ ̄*Y*). The physician faces a discrete choice problem: they must decide on incorporation status and, if incorporated, whether to constrain period 2 income to access the lower tax rate. This creates four scenarios for physicians who typically earn high income in period 1 (*wh*_1_ ≥ ̄*Y*):

##### Scenario 1: No Incorporation, High Period 2 Income The physician chooses *h*_2_ such that

*wh*_2_ ≥ ̄*Y*, facing *τ_p_* = *τ_H_* in both periods:

**Period 1 labour supply**:

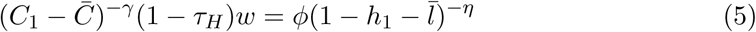

**Period 2 labour supply:**

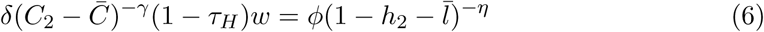

**Savings decision:**

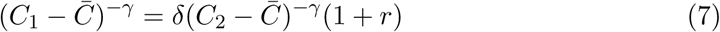

##### Scenario 2: No Incorporation, Low Period 2 Income The physician chooses *h*_2_ such that

*wh*_2_ *< ̄Y*, facing *τ_p_* = *τ_H_* in period 1 and *τ_p_* = *τ_L_* in period 2:

**Period 1 labour supply:**

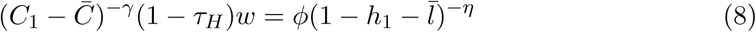

**Period 2 labour supply:**

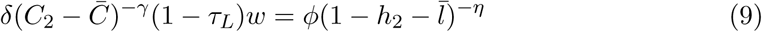

**Savings decision:**

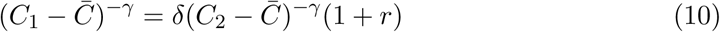

**Plus the constraint:** *wh*_2_ ≤ ̄*Y*

Scenario 3: Incorporation, High Period 2 Income The physician incorporates and chooses *h*_2_ such that *wh*_2_ + (1 + *r*)*S* ≥ ̄*Y*, facing *τ_p_* = *τ_H_*in both periods:

**Period 1 labour supply:**

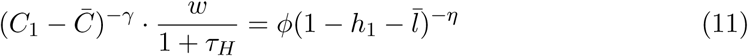

**Period 2 labour supply:**

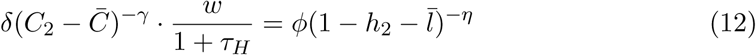

**Savings decision:**

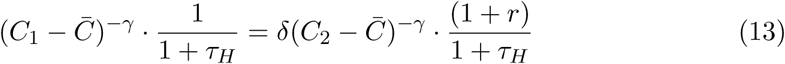

Scenario 4: Incorporation, Low Period 2 Income The physician incorporates and chooses *h*_2_ such that *wh*_2_ + (1 + *r*)*S < ̄Y*, facing *τ_p_* = *τ_H_*in period 1 and *τ_p_* = *τ_L_* in period 2: **Period 1 labour supply:**

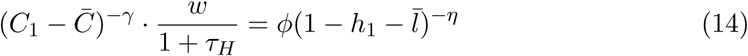

**Period 2 labour supply:**

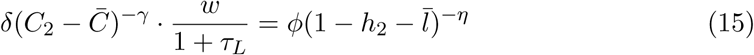

**Savings decision:**

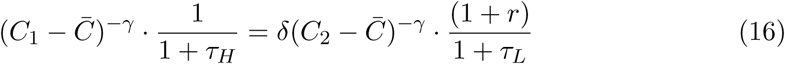

**Plus the constraint:** *wh*_2_ + (1 + *r*)*S* ≤ ̄*Y*

#### B.1.4 Dominance of Incorporation

We now establish that incorporation weakly dominates non-incorporation for all physician types. The key insight is that, under the assumption that incorporation is costless, incorporation provides two fundamental advantages: the ability to save from pre-tax income and enhanced flexibility in tax timing strategies. With Stone–Geary preferences, incorporation also enables agents to meet subsistence needs with less labour effort, which can trigger discontinuous changes in labour supply, such as early retirement.

Direct Comparisons: Same Period 2 Tax Strategy

**Proposition 1:** Scenario 3 (incorporation, high period 2 income) dominates Scenario 1 (no incorporation, high period 2 income).

*Proof:* Both scenarios face the same tax rates (*τ_H_* in both periods), but differ in the effective cost of saving. Without incorporation, each dollar saved requires sacrificing 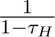 dollars of period 1 consumption, since saving must be made from after-tax income. With incorporation, each dollar saved requires sacrificing only one dollar of pre-tax income. This allows incorporated physicians to achieve the same post-subsistence consumption profile with less work effort, or alternatively, to achieve higher utility (more leisure or more saving) conditional on meeting the same subsistence thresholds.

**Proposition 2:** Scenario 4 (incorporation, low period 2 income) dominates Scenario 2 (no incorporation, low period 2 income).

*Proof:* Both scenarios achieve the low tax rate *τ_L_* in period 2, but incorporation provides the same pre-tax saving advantage as in Proposition 1. In addition, incorporated physicians have more flexibility in satisfying the period 2 income constraint, as they can jointly adjust both *h*_2_ and *S* to ensure *wh*_2_ + (1 + *r*)*S < ̄Y*, whereas unincorporated physicians can only adjust *h*_2_ to satisfy *wh*_2_ *< ̄Y* . With Stone–Geary preferences, this flexibility is particularly valuable: incorporation may allow the physician to meet the consumption subsistence level ̄*C* in period 2 with a smaller *h*_2_, potentially triggering a full exit from the labour force (*h*_2_ = 0) if leisure preferences are strong.

Cross-Comparisons: Different Period 2 Tax Strategies

**Proposition 3:** The best available strategy under incorporation dominates the best available strategy without incorporation.

*Proof:* An unincorporated physician chooses between a high-income strategy (Scenario 1) and a low-income strategy (Scenario 2), selecting the one that yields higher utility: max{*U*_1_*, U*_2_}. Similarly, an incorporated physician chooses between the incorporated analogues: Scenario 3 (high income) and Scenario 4 (low income), yielding utility max{*U*_3_*, U*_4_}. From Propositions 1 and 2, we know that *U*_3_ ≥ *U*_1_ and *U*_4_ ≥ *U*_2_. Therefore,

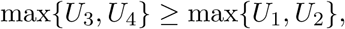

and incorporation weakly dominates non-incorporation for all physicians.

Economic Intuition The dominance result stems from incorporation expanding physicians’ choice sets along two dimensions. First, incorporation allows saving from pre-tax rather than after-tax income, effectively increasing the return to deferred consumption. Second, incorporation provides enhanced flexibility in period 2 tax management, as physicians can control their taxable income through both labour supply (*h*_2_) and accumulated savings (*S*), rather than labour supply alone.

With Stone–Geary preferences, these advantages are particularly salient for physicians near their subsistence thresholds. The marginal utility of consumption increases sharply as *C_t_* approaches ̄*C*, and the disutility of labour increases sharply as leisure falls below 1 − ̄*l*. This structure introduces kink points in behavior: physicians may exhibit smooth responses when far from these thresholds but sudden, discontinuous adjustments in labour supply, including abrupt retirement decisions occur once thresholds are crossed. Incorporation can trigger such threshold-crossing behavior by improving the feasibility of meeting minimum consumption needs with less labour effort, especially in period 2.

Implications for Labour Supply and Savings

**Period 1 Labor Supply (***h*_1_): The effect of incorporation on period 1 labour supply is ambiguous due to competing income and substitution effects. On one hand, the increased return to saving creates a substitution effect that encourages greater work effort in period 1 to fund higher consumption or earlier retirement. On the other hand, incorporation improves the overall budget constraint, making the physician effectively wealthier and incentivizing more leisure. The strength of these forces depends on how close the physician is to the consumption threshold ̄*C*. Those near the margin may exhibit sharp behavioural responses to small changes in return to saving, while those further away respond more smoothly.

**Savings (***S*): Incorporation strengthens the incentive to save by allowing physicians to accumulate savings from pre-tax income. This increases the effective return to deferred consumption from (1 + *r*) to 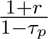 in after-tax terms. With Stone–Geary preferences, agents place especially high value on smoothing consumption above the subsistence level ̄*C*. The ability to defer tax and achieve smoother consumption across periods increases utility even if total consumption does not rise, and reinforces the motive to save. This is particularly the case for those who anticipate reduced labour supply in period 2.

**Period 2 Labor Supply (***h*_2_): Incorporation can lead to substantial reductions in period 2 labour supply, including full retirement, when the improved return to saving allows the physician to meet subsistence consumption needs without additional work. The presence of a minimum consumption threshold and steep marginal disutility of labour below the leisure threshold (̄*l*) creates the possibility of corner solutions. If the physician’s accumulated savings and low effective tax rate (via strategic income timing) are sufficient to achieve *C*_2_ ≥ ̄*C*, then the optimal response may be to set *h*_2_ = 0. Thus, the model predicts extensive-margin behavioural responses, such as abrupt retirements, that occur as a result of small improvements in tax efficiency.

The model yields several empirically testable implications. First, the effect of allowing incorporation on period 1 labor supply is theoretically ambiguous: while higher effective returns to savings generate a substitution effect that may increase work effort, the resulting increase in lifetime wealth also produces an income effect that may reduce effort. Empirically, both positive and negative intensive margin supply responses are feasible. The model also implies that labor supply in period 2 is theoretically ambiguous. While some physicians may continue working and smooth their consumption by drawing salary from their corporation, many may strategically reduce their period 2 income to fall below the higher personal tax threshold.

However, incorporation creates strong incentives for early retirement by enabling physicians to defer tax on savings and smooth consumption in retirement. This is both because older physicians may have lower discount factors and because they may be closer to their subsistence level of ̄*C* due to previous saving. Our model implies that, intensive margin ambiguity notwithstanding, we should observe a decline in the extensive margin supply of older physicians, particularly those near or past traditional retirement ages (e.g., 60+), shortly after incorporation is allowed. As well, because incorporation facilitates retirement planning over a physician’s entire career, the model also predicts that long-run extensive margin labor supply will decline, as successive cohorts retire earlier than they would have otherwise due to higher accumulated savings.

Finally, since only physicians with sufficient earnings can meaningfully benefit from tax deferral, we expect heterogeneous responses by specialty and income, with high-earning physicians (e.g., surgical specialists) showing stronger labor supply reductions. Finally, incorporation benefits are larger for those with longer career horizons and accumulated savings, so we also expect stronger responses among more experienced physicians, as measured by years since graduation. These implications guide our empirical analysis of both short-run and long-run changes in labor supply at the intensive and extensive margins, with a particular focus on age, graduation cohort, and specialty heterogeneity.

